# Mindfulness-Based Stress Reduction for Generalized Anxiety Disorder: Systematic Review and Meta-Analysis

**DOI:** 10.64898/2026.10.21.26366048

**Authors:** Vinicius V. Neves, Nana Amma O. O. Larbi, Rebecca L. Acabchuk, Laura N. Haygood, Vedhalakshmi Rajasankar, Simon B. Goldberg, Shufang Sun, Elizabeth A. Hoge, Judson A. Brewer, Chen Li, Blair T. Johnson, Eric B. Loucks

## Abstract

**Objective:** To evaluate the efficacy and safety of Mindfulness-Based Stress Reduction (MBSR) for anxiety symptoms in adults with generalized anxiety disorder (GAD).

**Design:** Systematic review and meta-analysis.

**Data Sources:** CINAHL, Embase, MEDLINE, PsycINFO, Scopus, SocINDEX, ERIC (EBSCO), and ProQuest Dissertations & Theses Global were searched from inception to October 2025; identified trial protocols were hand-searched for subsequent reports through February 2026.

**Eligibility criteria for selecting studies:** Randomized, non-randomized, and single-arm trials evaluating the effects of MBSR on anxiety in adults with GAD.

**Data extraction and synthesis:** Two authors independently screened records, extracted data, and assessed risk of bias. The primary outcome was anxiety symptoms; the secondary outcome was adverse events. Random-effects meta-analyses were conducted, and certainty of evidence was assessed.

**Results:** Of 19 included reports (16 independent samples), 8 randomized controlled trials (*n*=1,012) were included in the primary meta-analysis. MBSR significantly reduced anxiety symptoms vs. waitlist (Hedges’ *g*=−0.88; 95% CI −1.57 to −0.19) and general active comparators (*g*=−0.85; 95% CI −1.15 to −0.54), and showed no significant difference vs. escitalopram in two linked trials (*g*=0.09; 95% CI −0.11 to 0.29), with fewer adverse events in the in-person trial, the only one reporting them (15% vs. 79%; number needed to treat to avoid harm=2). GRADE certainty of evidence was high for the escitalopram comparison and very low for waitlist.

**Conclusions:** MBSR is a well-tolerated, nonpharmacologic treatment option for adults with GAD. Anxiety reductions were similar to escitalopram, with fewer adverse events; independent replication is needed. MBSR may be a viable option for patients who prefer non-pharmacological management or who cannot tolerate medication.

**Systematic review registration:** PROSPERO CRD420251140213.

**SUMMARY BOX:** *What is already known on this topic:*

- Generalized anxiety disorder (GAD) is common and disabling, and pharmacotherapy is frequently discontinued because of adverse effects
- Previous reviews of mindfulness-based interventions for anxiety pooled different programs and diagnoses, with inconsistent findings
- Recent noninferiority trials comparing Mindfulness-Based Stress Reduction (MBSR) with escitalopram had not been synthesized

*What this study adds:*

- MBSR reduced anxiety symptoms compared with waitlist and general active comparators, although certainty of evidence for the waitlist comparison was very low
- In two linked trials, anxiety reductions with MBSR were similar to those with escitalopram (high certainty evidence; g=0.09, 95% CI −0.11 to 0.29), with fewer adverse events in the in-person trial (15% v 79%; number needed to treat to avoid harm 2)
- Independent replication and head-to-head trials against cognitive behavioral therapy are needed

## INTRODUCTION

Generalized anxiety disorder (GAD) is among the most common anxiety disorders worldwide, causing psychological and physical symptoms that severely impair quality of life, productivity, and overall functioning.^1^ ^2^ A nationally representative survey of 43,000 U.S. adults suggests that GAD often causes functional impairment comparable to or exceeding that of major depressive disorder.^3–5^ Longitudinal data further indicate that changes in GAD symptoms are a significant predictor of functional impairment, even after controlling for comorbidities.^6^ Consequently, GAD carries a substantial economic burden, driving high healthcare costs and significant work impairment.^7–11^ The disorder’s chronic course and low remission rates compound this strain into a sustained societal impact.^7^ Although current treatments reduce symptoms for many patients, clinical responses are often unsatisfactory.^12^ Pharmacotherapy is frequently discontinued because of adverse effects;^13^ ^14^ cognitive behavioral therapy provides durable benefits, but many other psychotherapies show inconsistent efficacy.^15^ Moreover, many patients avoid traditional care and prefer to manage symptoms independently, potentially reflecting a desire for greater autonomy and self-management, or concerns about the stigma associated with mental health services;^16^ ^17^ these patterns highlight the need for effective, better-tolerated, skill-building alternatives.^12^ ^18^

Among mindfulness-based interventions (MBIs), Mindfulness-Based Stress Reduction (MBSR) is a standardized, group-based program that cultivates present-moment awareness and nonjudgmental acceptance through meditation, yoga, and mindfulness practices.^19^ MBSR shows promise for anxiety and has demonstrated benefits across mental health conditions including depression and post-traumatic stress disorder.^20^ ^21^ Although numerous systematic reviews have examined MBIs for anxiety broadly, specific evidence on MBSR for GAD remains fragmented. Prior reviews conflated MBSR with other MBIs^22–28^ or included non-GAD populations,^22–26^ ^29^ with inconsistent findings: some reported at least moderate benefits,^26^ ^27^ ^29^ whereas others reported weaker effects,^22^ ^25^ all of which varied considerably by control/comparator condition. Two recent reviews examined closely related but distinct questions: Williams et al. synthesized literature on MBIs broadly for GAD through November 2021,^27^ while Yumkhaibam et al. examined MBSR across multiple anxiety diagnoses through April 2024 and reported GAD findings narratively without quantitative synthesis.^24^ Both called for larger, higher-quality randomized-controlled trials (RCTs), and neither included recent landmark noninferiority trials,^30^ ^31^ underscoring the need for an updated synthesis.

Given the burden of GAD and the tendency of previous reviews to pool heterogeneous MBIs and populations, we focused specifically on MBSR for GAD to support more precise clinical conclusions. Accordingly, this systematic review and meta-analysis aimed to assess the effect of MBSR on anxiety symptoms in adults with GAD; compare MBSR with established treatments and other comparators; examine sources of variability in outcomes and the certainty of evidence; and compare adverse events between MBSR and comparators.

## METHODS

This review followed the principles of the Preferred Reporting Items for Systematic Reviews and Meta-Analyses (PRISMA) Statement^32^ and adhered to AMSTAR 2 methodological quality standard for systematic reviews.^33^ The study was preregistered with PROSPERO (CRD420251140213).

### Data Sources and Searches

A research librarian (LNH) drafted the Ovid Medline search around three concepts (MBSR, GAD, and validated anxiety outcome measures), combining title, abstract, and keyword text words with subject headings using Boolean operators. A second librarian peer reviewed the draft using the PRESS checklist.^34^ The finalized search was translated to 7 additional databases (CINAHL, Embase, APA PsycINFO, Scopus, SocINDEX, ERIC, and ProQuest Dissertations & Theses Global for gray literature) and run from inception to October 2025 (Appendix 2).

Trial protocols identified were hand-searched for subsequently published reports through February 2026. All records were exported to Covidence (Veritas Health Innovation), which removed duplicates.

### Study Selection

Studies were eligible for inclusion as follows: *(1) Study design:* RCTs, non-randomized controlled trials, and single-arm trials with independent samples, in any language, published or unpublished (see Appendix 1 for rationale). *(2) Participants:* adults (≥18 years) with GAD, diagnosed using the Diagnostic and Statistical Manual of Mental Disorders (DSM) or identified using validated instruments with substantial evidence supporting their accuracy for GAD. After reviewing the literature up to October 2025, our team concluded that Generalized Anxiety Disorder-7 (GAD-7) and the Hospital Anxiety and Depression Scale Anxiety subscale (HADS-A) were the primary self-report scales that fit these criteria^35^ ^36^ (see Appendix 1 for rationale). Reports with mixed populations were included if ≥50% of participants met GAD criteria; when this proportion was not reported, studies were included if the sample’s mean baseline score exceeded the established measurement tool-specific cutoff for GAD, a criterion intended to ensure clinically relevant symptom levels and to limit floor effects rather than to establish diagnostic status. *(3) Intervention group:* standard MBSR per the intervention manual,^37^ ^38^ or modified versions that adhered to specific fidelity criteria. Preregistered acceptable modifications were (a) reduction or removal of the all-day retreat, (b) shortening weekly sessions by no more than 30% of the standard 2.5 hours, or (c) reducing the eight-week curriculum by no more than 25% (excluding orientation). Interventions were required to be guided, group-based, and delivered synchronously in-person or online. Studies with modifications beyond these were excluded. *(4) Control/comparator:* unrestricted (RCTs prioritized in primary analysis). *(5) Outcomes:* anxiety symptoms, measured using validated clinician-rated or self-report tools.

NotebookLM (Google; now Gemini Notebook), an artificial intelligence (AI) powered tool, was utilized in accordance with PRISMA guidelines^32^ to automatically expedite the retrieval of specific data points regarding eligibility criteria from full-text articles (See Appendix 1 for prompt). All AI-identified data were manually verified against the source text by the reviewers to ensure accuracy.

### Data Extraction and Quality Assessment

A standardized data extraction form was developed based on the PRISMA 2020 statement^32^ and Cochrane reporting recommendations.^39^ Screening, extraction, and risk-of-bias assessment were each performed independently by two reviewers; critical quantitative outcome data were quadruple-coded. Interrater reliabilities were in the substantial agreement range (Appendix 1).^40^ ^41^ Discrepancies were resolved by discussion or by a third senior author (EBL, RLA, BTJ).

### Risk of Bias in Individual Studies

For RCTs, risk of bias was assessed with the Cochrane Risk-of-Bias 2 (RoB 2) tool;^42^ for Nonrandomized Studies of Interventions (NRSI), we used the Risk of Bias in NRSIs, Version 2 (ROBINS-I V2);^43^ and, for pre-post studies, we used the Quality Assessment Tool for Before-after (Pre-Post) Studies with No Control Group.^44^ Traffic-light plots and summary charts were generated.

### Data Synthesis and Analysis

#### Primary and Secondary Analyses

Primary analyses evaluated shorter-term MBSR efficacy in RCTs across three preregistered comparator strata: inactive (e.g., waitlist), general active (e.g., stress management education [SME], active treatment as usual [TAU]), and first-line anxiety pharmacotherapy. Secondary analyses examined treatment maintenance at longer-term follow-up in RCTs and assessed pre- to post-treatment changes (within-group effects) across RCTs, non-RCTs and single-arm trials.

#### Unit of Analysis

Consistent with standard meta-analytic practice, the independent sample (study), rather than the published report, served as the unit of analysis.^32^ ^45^ ^46^ Where multiple reports drew on the same sample (i.e., secondary analysis of a parent RCT), they were linked and treated as a single study by prioritizing the parent study’s outcomes to preserve independence and avoid double-counting.

#### Effect size calculations

All effect sizes were expressed as standardized mean differences with 95% CIs, incorporating a small-sample correction; negative values indicate a reduction in anxiety symptoms. For between-group (RCT) comparisons, the effect size was the standardized difference between arms (MBSR minus control) at post-treatment minus the corresponding standardized difference at baseline, removing any pre-existing between-arm imbalance. For within-group analyses, the pre–post change was standardized by the baseline standard deviation. Appendix 1 gives notation, formulae, and the source estimators.^47^ ^48^

One effect size was analyzed per study. When reports used multiple instruments, effect sizes were calculated from the highest-ranked tool in a preregistered clinical-validity hierarchy (Appendix 1), so that each study contributed a single independent estimate. For each arm and timepoint, means, standard deviations, and sample sizes were extracted.

#### Data Synthesis and Heterogeneity

Quantitative data were pooled using a random-effects model, as clinical and methodological diversity was anticipated across studies. To ensure the robustness of the synthesis, meta-analysis was performed only when *k*≥2 studies were available for a specific design or comparison type. Heterogeneity was assessed using the *I*^2^ statistic, Cochran’s *Q* test, and *τ*^2^, alongside visual inspection of forest plots. Where notable heterogeneity was detected, potential sources were explored using subgroup analysis or meta-regression. Between- and within-group comparisons were analyzed separately using the rma function of the metafor package (version 4.8.0) in R version 4.5.1.^47^ ^49^

#### Moderator Analyses

Moderator analyses were conducted to inform future trial design and explore heterogeneity sources, following criteria for assessing moderators in systematic reviews.^50^ All moderator variables listed below were prespecified in the protocol. Following Fu et al.,^51^ k≥4 studies per subcategory is recommended for categorical moderator tests; given the limited number of eligible trials, we adopted a threshold of k≥2 studies per level, and all moderator tests are accordingly hypothesis-generating rather than confirmatory. Prespecified moderators that did not meet this threshold were not tested. Moderator variables spanned three domains: methodological (study design, control condition type, risk of bias, conflict of interest disclosures, and eligibility and outcome assessment method), intervention (delivery format and MBSR curriculum adherence), and participant characteristics (study population, age, and gender).

### Publication Bias

To triangulate evidence across multiple approaches, small-study effects were assessed via funnel plot inspection and, when *k*≥10 studies were available (per the preregistration), Egger’s regression test. Trim-and-fill analyses were conducted to estimate potential missing studies, and sensitivity analyses evaluated the robustness of findings to these biases.

### Certainty of Evidence

The certainty of the evidence for the primary outcome was evaluated using the Grading of Recommendations Assessment, Development and Evaluation (GRADE) system.^52^ Evidence was categorized as “high,” “moderate,” “low,” or “very low” based on limitations in five domains: risk of bias, inconsistency, indirectness, imprecision, and publication bias.^52^ Assessments were conducted independently by two scientists, with discrepancies resolved through discussion with a third senior scientist. Because GRADE ratings apply to a specific comparison, certainty was rated only where pooled studies shared the same comparator type and timepoint.

### Patient and public involvement

The review question was prioritized by a formal stakeholder advisory panel that included patients, clinicians, health insurers, health care administrators, and policymakers, convened through an integrated knowledge translation approach.^53^ ^54^ The panel identified evaluation of MBSR for specific mental health conditions, particularly anxiety, as a critical evidence need, and identified evidence on effectiveness and side effects as essential for coverage decisions.^54^

## RESULTS

### Study Selection

The search yielded 1,560 records, 566 of which were duplicates. Hence, 994 records were screened at the abstract and title level, and 214 reports were retrieved for full-text screening. Of these, 195 were excluded for not meeting the inclusion criteria. The most common reasons for exclusion were: (1) insufficient information to confirm that ≥50% of participants met GAD criteria (*k*=75); (2) modifications to MBSR exceeding those specified in the preregistered protocol (*k*=58); (3) low prevalence of GAD (*k*=32) (see Appendix 3 for study list; and **Figure S1** for details).

### Study Characteristics

The 16 included studies enrolled 1,321 participants, 1,059 of them in RCTs; 2 studies had fewer than 30 participants. Participants’ mean age was 43 years, and 64% were female. Eligibility was established by validated scale cutoffs in 12 studies and by the Structured Clinical Interview for DSM^55^ or the Mini-International Neuropsychiatric Interview^56^ in four. **Table 1** summarizes all 19 reports, which represent these 16 independent samples/studies; three reports (Hoge^57^ ^58^; Hu^59^) are subsamples or secondary analyses of parent trials also included, and studies are the unit of analysis throughout.

**Table 1:**
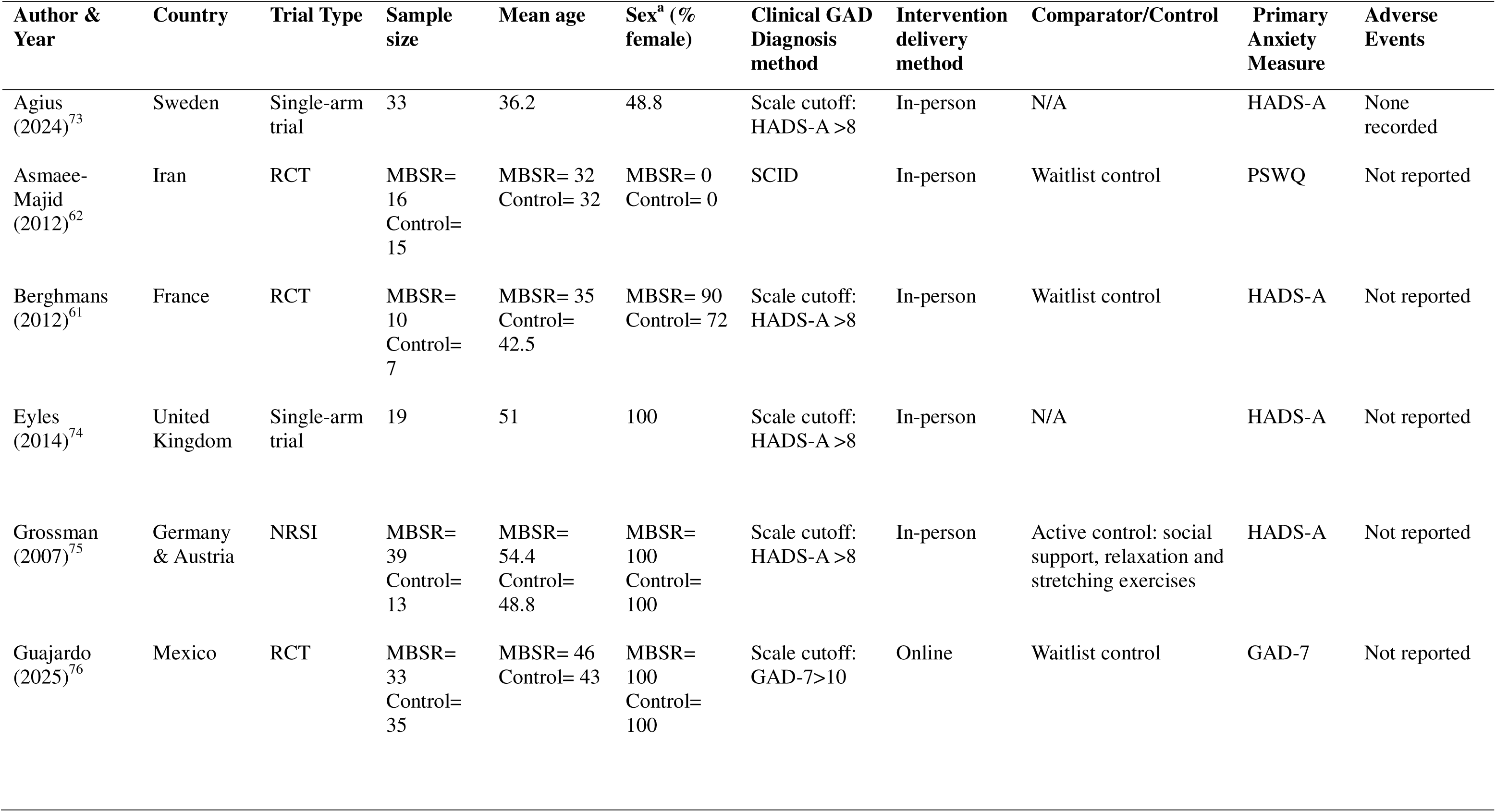
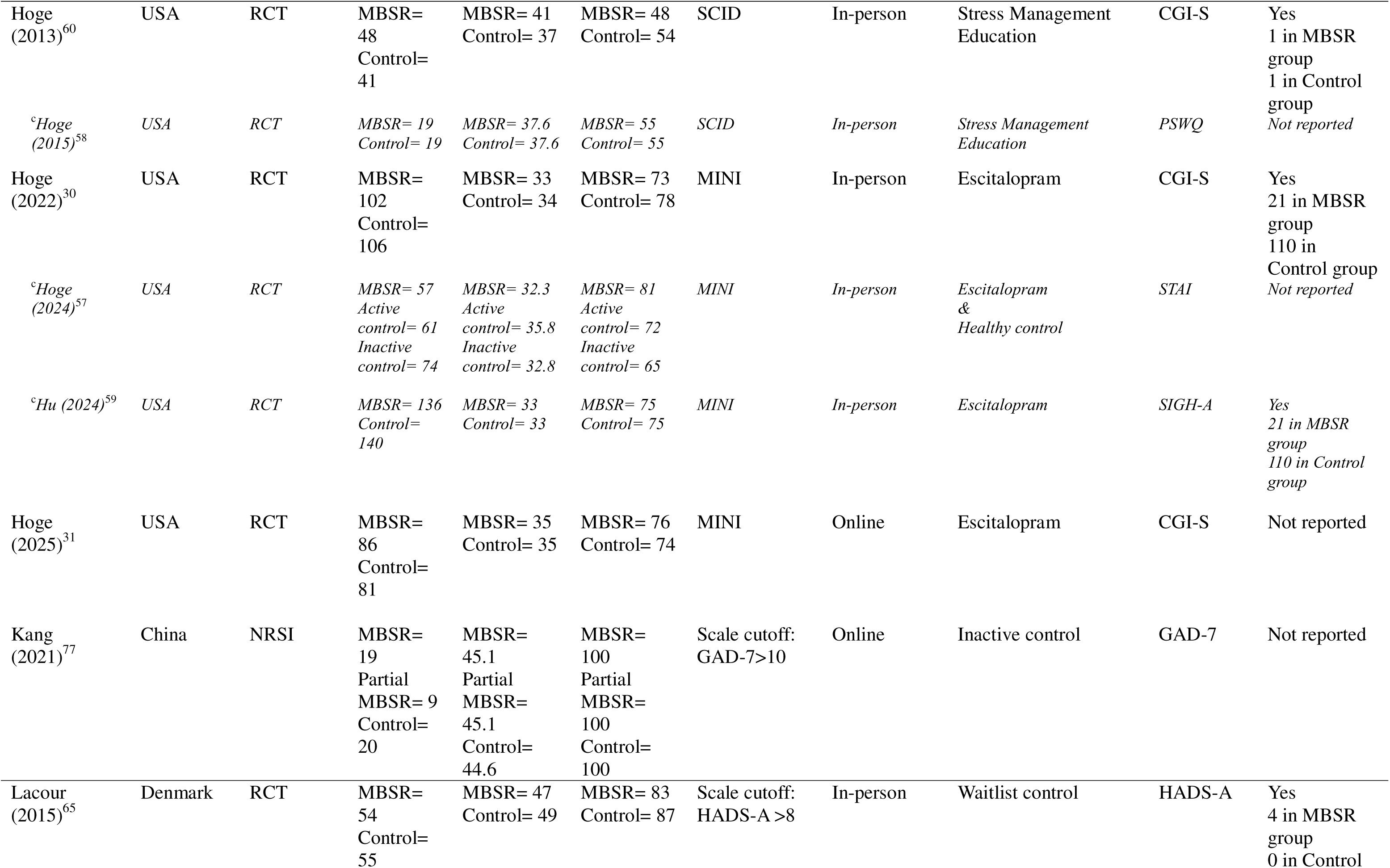
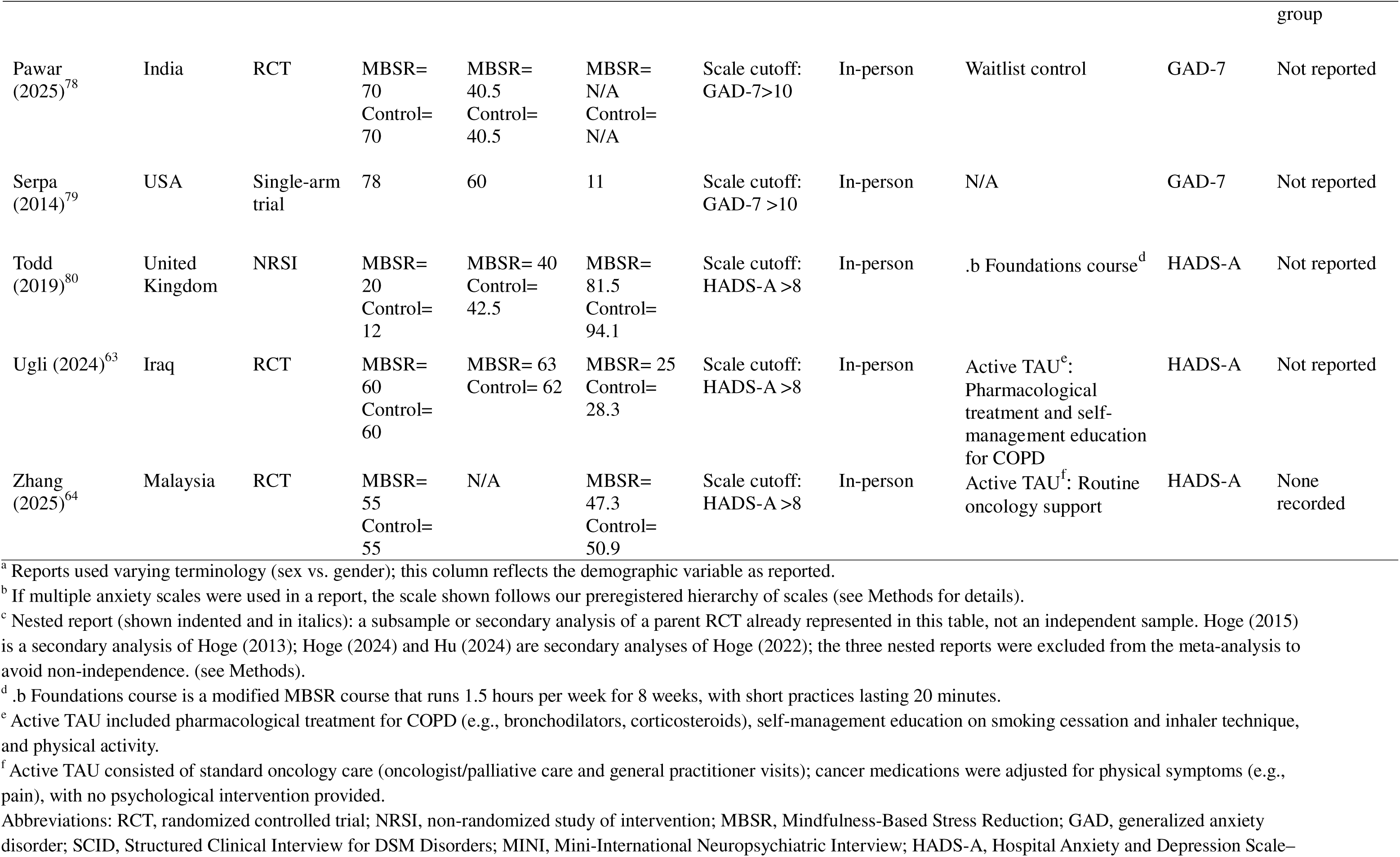
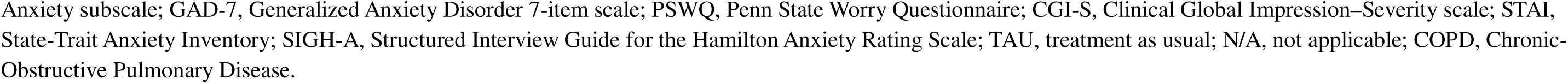
Characteristics of Included Reports/Studies.

Ten of the 16 studies (62%) were RCTs. Studies were conducted in North America (*k*=5), Europe (*k*=6), and Asia (*k*=5), and most interventions were delivered in person (*k*=13), with three using online platforms. The most common comparator on RCTs was waitlist control (*k*=5), followed by active treatment-as-usual (*k*=2), pharmacotherapy (escitalopram; *k*=2), and SME (*k*=1). Table S2 reports funding sources.

### Overall Quantitative Results

All 19 reports were reviewed narratively, but not all contributed to the meta-analyses. Three reports^57–59^ were not pooled separately, as their participants are represented in the parent trials.^30 60^ Two further studies could not be pooled: for one, the statistics required to calculate its effect’s standard deviation were unavailable, despite attempts to contact the authors;^61^ the other was conservatively withheld after a data integrity check revealed unusually large effect sizes (biased towards larger effects of MBSR) and discrepancies across calculation methods (Appendix 4).^62^ Fourteen studies therefore contributed effect sizes.

#### Between-Group Comparisons

##### Immediate Post-Treatment

**Figure 1** shows the primary analyses of 8 RCTs comparing MBSR to control conditions, stratified by comparator type. Compared with waitlist (*k*=3), MBSR showed a large, statistically significant superior effect (*g*=−0.88; 95% CI −1.57 to −0.19; *P*=0.013), with substantial heterogeneity (*I*^2^=89.0%; τ^2^=0.33). Compared with general active comparators (SME, TAU; *k*=3), MBSR again showed a significant effect (*g*=−0.85; 95% CI −1.15 to −0.54; *P*<0.001), with moderate, non-significant heterogeneity (*I*^2^=46.2%; τ²=0.03). Two trials used condition-specific treatment-as-usual [COPD,^63^ cancer^64^], classified as active because participants received ongoing disease management (**Table 1**) that could improve anxiety symptoms rather than no care. These comparators differed from MBSR in content and were not designed to match it for attention, expectancy, or treatment intensity; SME, the only attention-matched comparator, showed the smallest effect in this stratum (**Figure 1**). A sensitivity analysis reclassifying both as inactive did not markedly alter the inactive comparator estimate (*g*=−0.93; 95% CI −1.31 to −0.54). Noninferiority trials (*k*=2) compared MBSR to first-line GAD pharmacotherapy escitalopram. This comparative efficacy meta-analysis found no significant difference between the two treatments (*g*=0.09; 95% CI −0.11 to 0.29; *P*=0.40), and no detectable heterogeneity (*I*^2^=0%; τ²=0.00).

**Figure 1.**
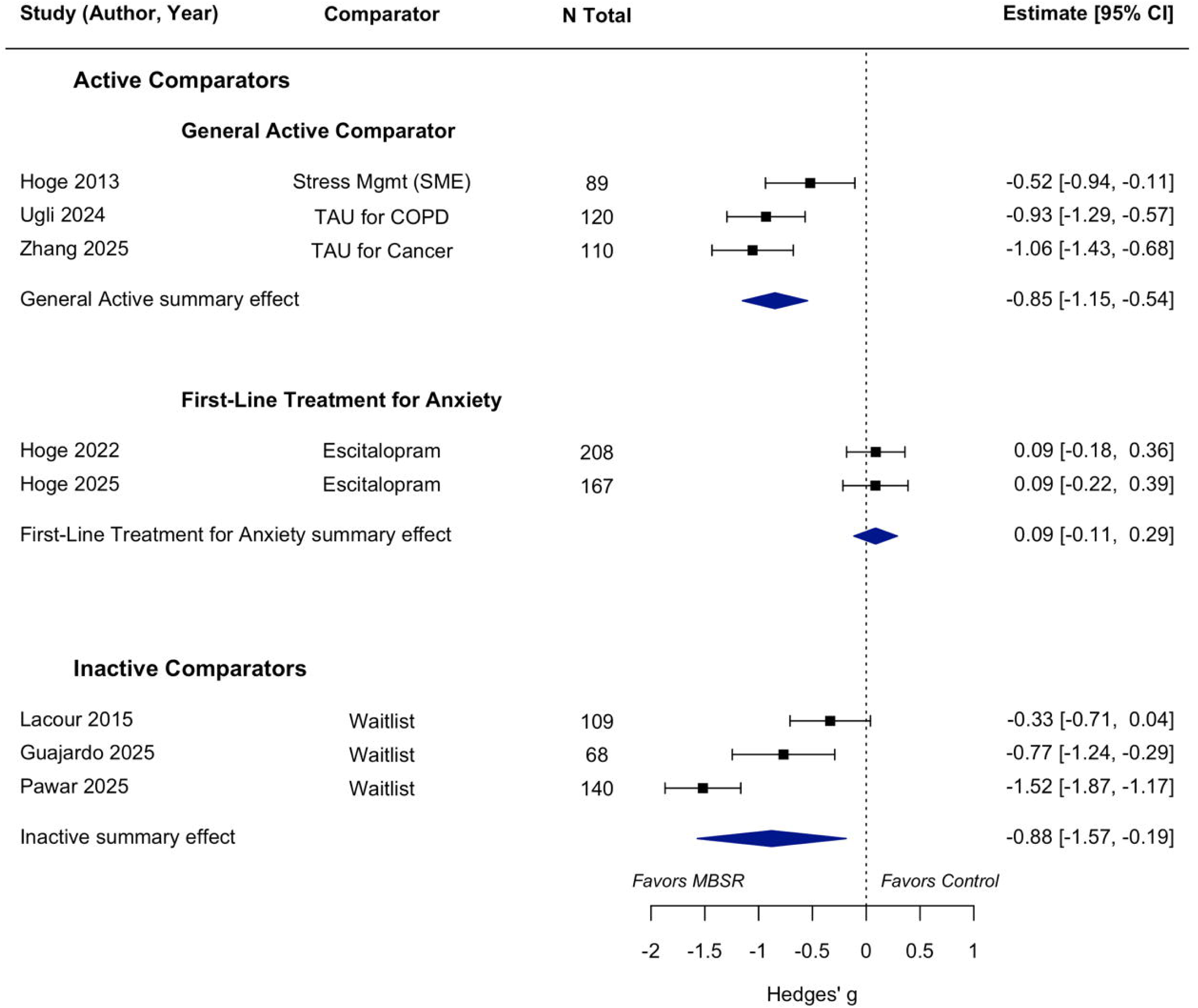
Forest plot of between-group differences in anxiety symptoms immediately after intervention, expressed as Hedges’ g. Change scores (post-treatment minus baseline) for the MBSR and control arms, standardized by the pooled SD and corrected for small-sample bias; pooled using a random-effects model and stratified by comparator type. All inactive controls were waitlist. Within active comparators, results are shown separately for general active comparators and for first-line pharmacotherapy (escitalopram in both trials). Diamonds=summary effects; negative values favor MBSR. CI=confidence interval; COPD=chronic obstructive pulmonary disease; MBSR=mindfulness-based stress reduction; SD=standard deviation; SME=stress management education; TAU=treatment as usual.

##### Longer-Term Follow-up

At longer-term follow-up (median 4 months, range 3–6 months; **Figure S2**), MBSR versus general active comparators (*k*=2) yielded a large, statistically significant effect favoring MBSR (*g*=−1.13; 95% CI −1.72 to −0.53), with substantial heterogeneity (*I*^2^=79.7%; τ²=0.15). Noninferiority trials comparing MBSR with escitalopram (*k*=2) showed no significant difference (*g*=0.10; 95% CI −0.11 to 0.32; *P*=0.33), with no detectable heterogeneity (*I*^2^=0%; τ²=0.00).

#### Within-Group Changes

##### Immediate Post-Treatment

The meta-analysis of within-group changes included 14 study arms assessing participants receiving MBSR. **Figure S3** shows the pooled results, indicating a large and statistically significant reduction in anxiety symptoms from baseline to immediate post-treatment (*g*=−1.06; 95% CI −1.36 to −0.75; *P*<0.001). High heterogeneity was observed (*I*^2^=93.0%; τ²=0.30). Results were consistent in direction across different individual study designs (**Figure S3**).

##### Longer-Term Follow-Up

**Figure S4** shows the durability of anxiety reduction, assessed across 9 study arms at longer-term follow-up (median 4 months, range 3–36). Each study contributed a single within-group standardized mean change from baseline to its longest available follow-up, pooled using a random-effects model. Improvements were sustained over time, with a large significant effect relative to baseline (*g*=−1.08; 95% CI −1.48 to −0.67; *P*<0.001). Heterogeneity remained high (*I*^2^=92.3%; τ²=0.34).

### Safety Profile and Adverse Events

Adverse events were documented and reported in 5 of 16 studies (31%); a quantitative synthesis was not conducted given substantial heterogeneity in how safety was monitored, ranging from passive surveillance to active clinician-led assessments. Across the three trials reporting specific occurrences,^30^ ^60^ ^65^ events in the MBSR arms were mild to moderate. Notably, in the largest trial comparing MBSR to escitalopram, which used active clinician-led monitoring, 21 participants (15% of group sample) in the MBSR group reported at least one study-related adverse event, compared to 110 participants (79% of group sample) in the escitalopram group.^30^ This 63 percentage-point absolute difference yields a number needed to treat (NNT) to avoid harm of 2, meaning 1 in every 2 patients treated with MBSR instead of escitalopram is spared at least one adverse event. No MBSR participants discontinued for adverse events, versus 8% (*n*=10) in the escitalopram arm. Appendix 6 provides study-level breakdowns and event-type frequencies.

### Moderator Analyses

Because comparator type, risk of bias, and outcome measure type were highly correlated across this small set of trials, their effects could not be disentangled; therefore all moderator results should be interpreted as exploratory. **Table S1** reports moderator analyses for both within-group and between-group comparisons. We prioritized the within-group moderator analyses because they included more study arms (*k*=14 vs. *k*=8) and were less susceptible to confounding by comparator type. Throughout, *B* represents the difference in Hedges’ *g* between moderator levels, with negative values indicating a greater reduction in anxiety symptoms. Three within-group moderators were statistically significant: a modified MBSR curriculum was associated with smaller pre-post reductions in anxiety symptoms than standard protocols (*B*=0.682; 95% CI 0.169 to 1.196; *P*=0.009); studies with scale-based eligibility and self-reported outcomes showed smaller reductions than those with structured-interview eligibility and clinician-rated outcomes (*B*=0.737; 95% CI 0.160 to 1.313; *P*=0.012); and synchronous online delivery with larger reductions than in-person delivery (*B*=−0.821; 95% CI −1.500 to −0.142; *P*=0.018).

### Risk of Bias Within and Across Studies

**Figure 2** shows ratings on the risk of bias of the included RCTs (*k*=10). Overall, most trials were assessed as having a high risk of bias (*k*=8), with only two independent samples/studies achieving a low-risk rating across all domains.^30^ ^31^ One additional study was classified as having some concerns.^60^ The high prevalence of bias was primarily driven by concerns in Domain 4 (measurement of the outcome, largely because several studies used self-reported anxiety scales among participants who were not blinded to assignment to the MBSR versus control condition), and Domain 5 (selection of the reported result), given the absence of pre-registered protocols or clinical trial registry entries in several studies, which raised concerns about selective outcome reporting across much of the sample.

**Figure 2.**
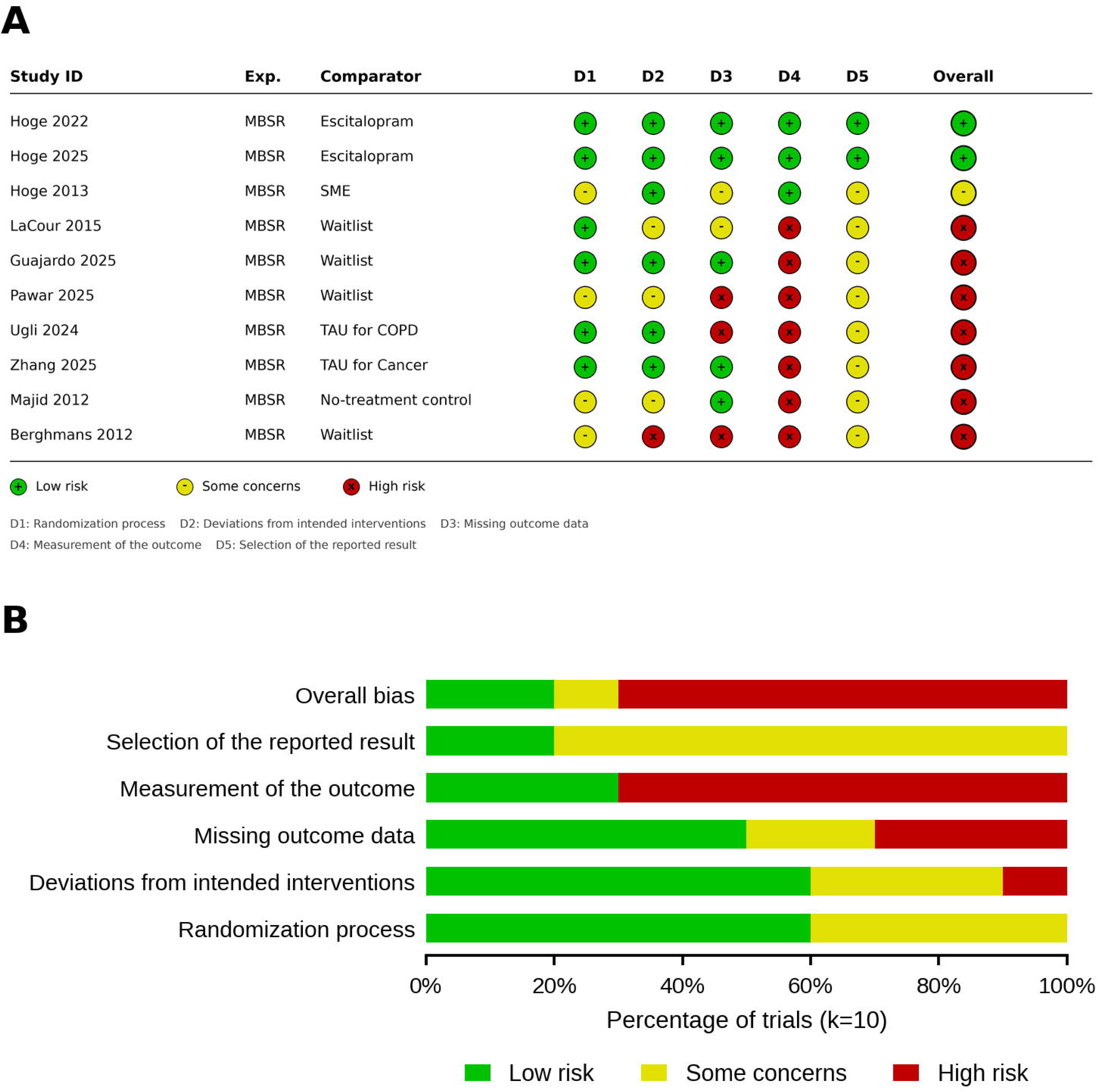
Risk of bias in the included randomized controlled trials (k=10). A, Cochrane Risk of Bias 2 judgments for each trial across five domains and overall. B, distribution of judgments within each domain, as a percentage of trials. D1=randomization process; D2=deviations from intended interventions; D3=missing outcome data; D4=measurement of the outcome; D5=selection of the reported result. MBSR=mindfulness-based stress reduction; SME=stress management education; TAU=treatment as usual.

Among the three non-RCTs, one was judged to be at medium risk of bias, while the other two were at serious and critical risk of bias. Lastly, all three single-arm trials were judged to be of fair quality.

Gray literature search identified no unpublished studies meeting the inclusion criteria. Visual inspection of the funnel plots (**Figure S5**) suggested small-studies effects at the between-group immediate post-intervention endpoint, prompting a post-hoc Egger’s test as a deviation from the pre-registered *k*≥10 threshold (*k*=8). This test was non-significant (*t*=−1.76, *df*=6, *P*=0.13) and trim-and-fill imputed no additional studies. Within-group funnel plots were approximately symmetrical; Egger’s test was statistically significant (*t*=−2.29, *df*=12, *P*=0.041), although trim-and-fill imputed no additional studies. Given substantial heterogeneity linked to differences in comparator type and study design (**Table S1**), these results are best interpreted as exploratory. Yet, as trim-and-fill imputed no studies in either analysis, and a robust gray literature search was conducted, any possible small-study effects are unlikely to have meaningfully altered reported estimates.

### Certainty of Evidence

GRADE was applied to the three comparisons in which pooled studies shared the same comparator and timepoint (**Table 2**): MBSR versus escitalopram at short-term and long-term follow-up, and MBSR versus waitlist at short-term follow-up. The general active stratum was not graded because its studies used different comparators (SME, COPD-specific TAU, and cancer-specific TAU). Both escitalopram comparisons were rated as high certainty, whereas the certainty of evidence for MBSR versus waitlist on short-term anxiety outcomes was rated as very low.

**Table 2:** GRADE Summary of Findings — MBSR for Anxiety in Generalized Anxiety Disorder (short-term = immediately post-intervention; long-term = 4 months post-intervention)

| Comparison | *k* (*n*) | Hedges' *g*<br>(95% CI) | Risk of Bias | Inconsistency | Indirectness | Imprecision | Publication<br>Bias | GRADE of<br>Certainty |
| --- | --- | --- | --- | --- | --- | --- | --- | --- |
| MBSR vs.<br>Escitalopram<br>(Short-term) | 2<br>(375) | 0.09 (−0.11 to<br>0.29) | Not Serious | Not Serious | Not Serious | Not Serious | Not Serious | ⊕⊕⊕⊕<br>High |
| MBSR vs.<br>Escitalopram<br>(Longer-term) | 2<br>(343) | 0.10 (−0.11 to<br>0.32) | Not Serious | Not Serious | Not Serious | Not Serious | Not Serious | ⊕⊕⊕⊕<br>High |
| MBSR vs.<br>Waitlist (Short-term) | 3<br>(317) | −0.88 (−1.57<br>to −0.19) | Very<br>Serious<sup>1</sup> | Serious<sup>2</sup> | Serious<sup>3</sup> | Serious<sup>4</sup> | Not Serious | ⊕⊖⊖⊖<br>Very Low |

*k* = number of studies; *n* = total participants; 95% CI = 95% confidence interval; ⊕ = certainty present; ⊖ = certainty reduced.
<sup>1</sup> Very Serious (Risk of Bias): Downgraded 2 levels. All *k*=3 included studies were assessed at high risk of bias overall, with high risk in outcome measurement (D4) due to absence of blinding combined with self-report scales, and additional high risk in missing outcome data (D3).
<sup>2</sup> Serious (Inconsistency): Downgraded 1 level. Significant heterogeneity was detected ( $I^2 = 89.0\%$ ), with point estimates varying widely in magnitude and non-overlapping confidence intervals.
<sup>3</sup> Serious (Indirectness): Downgraded 1 level due to population skew. One study was 100% female and others exceeded 83% female, limiting applicability to the broader GAD population of males.
<sup>4</sup> Serious (Imprecision): Downgraded 1 level. While the direction of effect consistently favored MBSR, the 95% confidence interval (−1.57 to −0.19) was very wide, spanning from a small to a very large clinical effect, precluding conclusions about the magnitude of benefit.

## DISCUSSION

This systematic review found that MBSR produced a moderate-to-large reduction in anxiety symptoms among adults with GAD immediately post-intervention, with significant effects compared to both waitlist and general active comparators such as SME and TAU. Meta-analysis of two noninferiority trials found no significant differences between MBSR and escitalopram, a first-line pharmacotherapy, post-intervention and at 4-month follow-up, with no detectable heterogeneity. The two trials were linked: the second was a pandemic-era videoconference cohort conducted under the same registration, protocol, and investigative team, with newly recruited participants and some changes in instructors. Although this was not a fully independent replication, and the videoconference cohort alone did not establish noninferiority, the pooled estimate (*g*=0.09; 95% CI −0.11 to 0.29) lay within the trials’ prespecified noninferiority margin (0.495 CGI-S points, approximately *g*=0.46). The in-person trial, the only one to report adverse events, found substantially fewer adverse events with MBSR than with escitalopram.

These findings advance earlier evidence syntheses that examined MBIs more broadly or across mixed anxiety populations.^22–29^ By focusing on standardized MBSR in predominantly GAD populations and applying GRADE, this review provides directly clinically actionable evidence for weighing MBSR against established treatments.

MBSR may be a reasonable treatment option for patients who prefer a nonpharmacologic approach, cannot tolerate medication, or are uncomfortable seeking help in mental health service settings. Its tolerability may be particularly relevant given the adverse effects associated with escitalopram, such as insomnia, sexual dysfunction, and fatigue.^13^ ^14^ In the head-to-head trial with active safety monitoring, adverse events occurred in 15% of participants receiving MBSR and 79% receiving escitalopram (NNT to avoid harm=2). No participant discontinued MBSR because of an adverse event, compared with 8% of those receiving escitalopram.^30^ ^31^ These findings support consideration of MBSR in clinical referral and reimbursement pathways, although treatment selection should also weigh patient preferences, availability, cost, and evidence certainty.

An additional implementation consideration is the time commitment MBSR requires, which typically encompasses an orientation session, eight weekly 2.5-hour group sessions, a 7.5-hour retreat, and approximately 45 minutes of daily home practice. Because it included only standardized or minimally adapted MBSR programs, this synthesis did not capture adaptations that may substantially reduce this burden, recognizing that the minimally adapted MBSR programs in this meta-analysis showed evidence of smaller effects than standardized MBSR. Regarding larger adaptations, some anxiety-specific programs have shortened group sessions, home practice, and retreats.^66^ These adaptations may be more acceptable to patients with GAD and could improve engagement and efficacy, but these possibilities cannot be evaluated in a review limited to standardized MBSR. Although abbreviated programs may no longer meet criteria for the “MBSR” designation,^54^ ^67^ ^68^ their shorter duration could facilitate implementation and reimbursement when delivered by or in partnership with licensed clinicians through group therapy or group medical visits.^54^ The acceptability, efficacy, and implementation of these adaptations warrant further research.

Ultimately, these findings provide comparative evidence that may inform payer coverage and reimbursement decisions. Under the GRADE framework, the evidence comparing MBSR with escitalopram was rated as high certainty and aligns with the Triple Aim framework utilized in healthcare systems: similar reductions in anxiety symptoms (*aim 1: improving population health*), fewer adverse events and adverse event-related discontinuations (*aim 2: improving patient experience*), and potential economic benefits *(aim 3: reducing per capita costs*).^69–71^ Supporting the latter, a companion economic-evaluation review found MBSR cost-savings relative to TAU across mental health populations.^72^

This review has several strengths, including its focus on GAD, inclusion of recent high-quality RCTs directly comparing standardized MBSR with a first-line pharmacotherapy, and systematic application of the GRADE framework. These strict eligibility criteria were important for a review intended to inform policy and yield more clinically applicable conclusions than those of previous syntheses pooling heterogeneous interventions or populations. However, this rigor also limited the sample: reports using adaptations of standard MBSR or lacking sufficient information to estimate whether at least 50% of participants had GAD were excluded. To maximize inclusion despite widespread underreporting, anxiety-scale cutoffs reported as accurate in Cochrane reviews were used to establish eligibility when diagnostic status was not reported.^35^ ^36^ Since self-report measures are less diagnostically precise than structured clinician interviews, this approach introduces uncertainty regarding participants’ diagnostic status and highlights a broader reporting gap in the field.

Additional limitations include the small number of trials that exclusively enrolled participants with GAD, frequent reliance on self-report outcomes at higher risk of bias, as well as substantial heterogeneity that could not be adequately explored through moderator analyses because of sample size restrictions. Comparison type, risk of bias, outcome assessment methods, and delivery format (all online programs ran during the pandemic) were highly correlated within this limited set of trials: all three trials with structured-interview eligibility and clinician-rated outcomes used active comparators,^30^ ^31^ ^60^ and two of these were the only studies judged to be at low risk of bias.^30^ ^31^ Stratification by comparator type, despite its apparent influence on effect size, left too few studies within each stratum (active comparators, *k*=3–5; inactive comparators, *k*=3) to estimate moderator effects reliably. Comparison type may therefore be an important source of between-study variation, but its influence—and that of the other potential moderators—should be interpreted cautiously. Safety evidence was also limited: adverse events were documented in only 5 of 16 studies, and the comparative estimate comes from the in-person trial, the only head-to-head trial reporting adverse events.

In conclusion, MBSR appears to be a well-tolerated, nonpharmacologic treatment for adults with GAD, with high-certainty evidence from 2 linked trials indicating anxiety outcomes similar to those of escitalopram and substantially fewer adverse events in the in-person trial. These findings support consideration of MBSR for patients who prefer nonpharmacologic management or cannot tolerate medication. Future RCTs should consider extended follow-up time periods, use structured diagnostic interviews to establish eligibility, report diagnosis-specific sample composition, and include blinded clinician-rated severity measures alongside patient-reported outcomes. Head-to-head trials against cognitive behavioral therapy, which provides durable benefits for GAD,^15^ and further pharmacotherapy comparisons are also needed; the only identified comparison with cognitive behavioral therapy tested an adapted MBSR program.^66^ Pragmatic trials should also evaluate implementation in routine care—including delivery through group therapy or group medical visits^54^—using standardized adverse-event ascertainment and formal cost-effectiveness analyses.

## Declarations

## Data Availability

The study protocol, statistical code, and dataset supporting these findings are available from the corresponding author upon reasonable request.

## Acknowledgment

The authors thank Amitojeroop Kaur, BA, University of Connecticut, whose honors thesis provided conceptual work on which this review was built. Her thesis informed the research question, eligibility criteria, and preliminary search strategy.

## Ethical approval

Not required.

## Transparency

The lead author (VVN) affirms that the manuscript is an honest, accurate, and transparent account of the study being reported; that no important aspects of the study have been omitted; and that any discrepancies from the study as planned (and, if relevant, registered) have been explained.

