## Appendix for "Mindfulness-Based Stress Reduction for Generalized Anxiety Disorder: Systematic Review and Meta-Analysis"

### Supplemental Material

#### Table of Contents

Appendix 1. Additional Methodological Details
Appendix 2. Full Search Strategy
Appendix 3. List of Excluded Trials and Reasons
Appendix 4. Exclusion of Study: Data Integrity Assessment
Appendix 5. Supplemental Figures and Tables
Figure S1. PRISMA Flowchart of Study Selection
Figure S2. Forest plot of longer-term between-group differences in anxiety symptoms
Figure S3. Within-group change from baseline to immediate post-treatment
Figure S4. Within-group change from baseline to longer-term follow-up
Figure S5. Funnel plots assessing small-studies effects
Table S1. Pre-registered Moderator Analysis
Table S2. Funding Sources for Included Studies
Appendix 6. Adverse Events: Study-Level Detail
References

### Appendix 1: Additional Methodological Details

#### Rationale for Expanding Inclusion Beyond Randomized-Controlled Trials

Non-randomized and single-arm designs were eligible for two reasons. First, while our primary outcome is a causal question, several secondary aims, including the moderator analyses, are correlational and do not require a randomized comparator to be informative; these reports were evaluated for methodological rigor using design-appropriate risk-of-bias tools, described below. Second, because RCT effects in this field are routinely heterogeneous and sensitive to control-condition characteristics, within-subject pre-to-post change was treated as a valuable complement to, though analyzed separately from, the RCT-based primary outcome.

#### Anxiety Self-report Validated Diagnostic Instruments Rationale

The GAD-7 is the most extensively validated and widely used screening tool for generalized anxiety disorder in adults. A recent Cochrane Review (1) reports a pooled sensitivity of 0.64 and specificity of 0.91 at the recommended diagnostic cutoff of ≥10. The area under the curve (AUC) of 0.86 demonstrates strong overall accuracy for detecting GAD. This interpretation is further supported by the U.S. Preventive Services Task Force Recommendation Statement (1,2), which identifies GAD-7 ≥10 as an empirically supported threshold for detecting clinically significant generalized anxiety disorder. Another Cochrane meta-analysis (3) demonstrates that the HADS-A subscale has comparable diagnostic performance to the GAD-7 for identifying generalized anxiety disorder. At the recommended cutoff of ≥8, HADS-A shows a sensitivity of 0.82 and specificity of 0.74, with an AUC of 0.82 for detecting GAD. Studies with mixed populations were included if ≥ 50% of participants meet GAD criteria. Those with <50% of participants with GAD were excluded. For self-report scale studies, some explicitly reported the proportion of participants scoring above the diagnostic cutoff on GAD-7 or HADS-A. For others, where this proportion was not provided, studies were included if the baseline mean score exceeded the established cutoffs (as outlined above), to ensure clinically relevant symptom levels and limit floor effects rather than to establish diagnostic status.

#### NotebookLM (Now Gemini Notebook) Rationale and Prompt to Support Data Extraction

Recent literature suggests that large language models can achieve screening sensitivity comparable to or exceeding human reviewers in systematic reviews, thereby streamlining the process and significantly reducing workload without compromising quality (4). The Notebook LM prompt used was as follows:

You are assisting with eligibility screening for a systematic review and meta-analysis of Mindfulness-Based Stress Reduction (MBSR) for generalized anxiety disorder (GAD). I will provide one study as a source and need you to extract only the information needed to confirm whether it meets our eligibility criteria. Use only what is stated in the source — do not infer, estimate, or calculate values that are not explicitly given. For every value, indicate where it appears (page, table, or figure) and quote the relevant text or table cell. Flag anything ambiguous or conflicting.

Extract:

1. GAD prevalence — how GAD diagnosis was established (e.g., structured clinical interview, DSM-5/DSM-IV criteria, screening-scale cut-off) and what proportion or number of the analyzed sample met criteria for GAD. If prevalence is not directly reported, extract the baseline score on any anxiety measure used, separately for each study group and the overall sample average, and name the measure(s), so this can be used as a proxy for ≥ 50% of participants meet GAD criteria.

2. MBSR protocol adherence — extract the following against our eligibility specification below:
- Session length (to check against the standard 2.5-hour session, reduced by no more than 30%)
- Total curriculum length in weeks (to check against the standard 8 weeks, reduced by no more than 25%, excluding any orientation session)
- Presence, reduction, or removal of the all-day retreat
- Whether the intervention was guided (instructor-led), group-based, and delivered synchronously (in-person or online)
- Any other stated deviation from the standard MBSR curriculum.

Present the result as a short summary per study, organized under these two headings, with page/table citations for each value.

#### Interrater Reliability

Screening: Cohen's κ = 0.629. Risk of bias (50 paired domain judgments, RoB 2): 70% exact agreement, linear-weighted κ = 0.62. Data extraction: 83% agreement across extracted cells. All discrepancies resolved by consensus.

#### Outcome Measurement Tool Preregistered Hierarchy for Effect Size Estimation

Following expert psychiatrist-scientist recommendation from co-author Elizabeth Hoge MD, anxiety measurement tools were prioritized to report on, ordered by importance, as follows:

1. Clinical Global Impression-Severity scale (CGI-S)
2. Structured Interview Guide for the Hamilton Anxiety Rating Scale (SIGH-A) or Hamilton Anxiety Rating Scale (HAM-A)
3. Penn State Worry Questionnaire (PSWQ)
4. Patient-Reported Outcomes Measurement Information System – Anxiety (PROMIS-A)
5. Beck Anxiety Inventory (BAI)

6. Generalized Anxiety Disorder-7 (GAD-7)
7. Hospital Anxiety and Depression Scale – Anxiety subscale (HADS-A)
8. State-Trait Anxiety Inventory (STAI)
9. Depression Anxiety Stress Scales-21 (DASS-21)

##### Effect Size Calculation Details

For each arm and timepoint, M, SD, and n denote the mean, standard deviation, and sample size. In the following paragraphs, subscripts "pre" and "post" denote baseline and post-treatment assessment. All effect sizes are standardized mean differences (SMDs) with Hedges' small-sample correction applied, reported with 95% CIs. Negative values indicate a reduction in anxiety symptoms.

For between-group analyses (RCTs): at each timepoint, the difference between arms (MBSR minus control) was standardized by the pooled standard deviation at that timepoint, and the baseline value was subtracted from the post-treatment value (Cohen's $d = d_{post} - d_{pre}$). This method ensures the resulting effect size reflects the true magnitude of change attributable to the intervention, independent of initial differences between the groups. Hedges' small-sample correction was then applied to the difference and its variance. Negative values indicate greater symptom reduction under MBSR.

For within-group analyses, pre-post change was standardized by the baseline standard deviation (Becker's *d*) (5), with Hedges' small-sample correction. Adjusted effect estimates controlling for confounding were not available for the single-arm and non-randomized studies, as these primary studies typically reported only unadjusted pre-post means and standard deviations; raw pre-post change was therefore used for all within-group analyses. Where the pre-post correlation needed to estimate variance was not reported, a value of 0.7 was assumed. Where standard deviations required for effect size calculation were not reported, study authors were contacted; if data could not be obtained, the study was excluded from the affected analysis.

#### Sensitivity Analyses

Two sensitivity analyses were conducted. First, to test the robustness of the active-comparator classification, the two trials using condition-specific treatment-as-usual (COPD, cancer) were reclassified as inactive comparators (6,7). Second, an influence analysis was conducted to assess the impact of a statistical outlier (8) on the pooled estimate and heterogeneity.

### Appendix 2: Full search strategy

*Databases:*
The following databases were searched: Ovid MEDLINE, Embase via Elsevier, SCOPUS, APA PsycINFO via EBSCO, CINAHL, SocINDEX via EBSCO, ProQuest's Dissertations & Theses Global.

We included papers written in all languages. Each database was searched from inception to October 2025.

The scientific research librarian conducted the searches, translated the search criteria for each database, uploaded resulting records into Covidence, and removed duplicate records.

*Search:*
Key concepts for search terms included:

1. Generalized Anxiety Disorder (e.g., "Generalized Anxiety Disorder," "GAD," "anxiety disorder")
2. MBSR, Mindfulness-Based Stress Reduction

A draft of a replicable search strategy for each individual database is below.

| | | |
|---|---|---|
| Database | Medline | |
| Platform | Ovid | |
| Date of last search | October 8, 2025 | |
| Number of results | 189 Results | |
| Notes | Ovid MEDLINE(R) ALL <1946 to October 07, 2025> | |
| 1 | Anxiety Disorders/ or "Generalized Anxiety Disorder"/ | 45269 |
| 2 | (GAD or Generalized Anxiet* or General* Anxiety Disorder* or Overanxious Disorder*).ti,ab,kf. | 24875 |
| 3 | ((PHQ* or "patient health questionnaire*" or "Clinical Global Impression*" or CGI* or SCID or "structured clinical interview" or "Depression Anxiety Stress Scale*" or DASS or "Hospital Anxiety and Depression Scale*" or HADS) and (anxi* or GAD)).ti,ab,kf. | 31811 |
| 4 | ("Beck anxiety inventor*" or BAI or "Hamilton Anxiety Rating" or HAM-A or "State-Trait Anxiety Inventory" or STAI | 23683 |

| | | |
|---|---|---|
| | or GAD-7 or "Liebowitz Social Anxiety Scale*" or LSAS* or "Penn State Worry Questionnaire*" or PSWQ).ti,ab,kf. | |
| 5 | or/1-4 | 101605 |
| 6 | "Mindfulness-Based Stress Reduction"/ | 1 |
| 7 | ("MBSR" or "mindfulness-based stress reduction" or "MBSR/MBCT").ti,ab,kf. | 1835 |
| 8 | (mindfulness-based adj3 stress).ti,ab,kf. | 1833 |
| 9 | mindfulness/ and stress reduction.ti,ab,kf. | 1071 |
| 10 | or/6-9 | 2070 |
| 11 | 5 and 10 | 189 |

| | | |
|---|---|---|
| Database | Embase.com | |
| Platform | Elsevier | |
| Date of last search | October 15, 2025 | |
| Number of results | 189 | |
| Notes | | |
| #1 | 'anxiety disorder'/de OR 'generalized anxiety disorder'/de | 132988 |
| #2 | gad:ti,ab,kw OR 'generalized anxiet*':ti,ab,kw OR 'general* anxiety disorder*':ti,ab,kw OR 'overanxious disorder*':ti,ab,kw | 37891 |
| #3 | (phq*:ti,ab,kw OR 'patient health questionnaire*':ti,ab,kw OR 'clinical global impression*':ti,ab,kw OR cgi*:ti,ab,kw OR scid:ti,ab,kw OR 'structured clinical interview':ti,ab,kw OR 'depression anxiety stress scale*':ti,ab,kw OR dass:ti,ab,kw OR 'hospital anxiety and depression scale*':ti,ab,kw OR hads:ti,ab,kw) AND (anxi*:ti,ab,kw OR gad:ti,ab,kw) | 52782 |
| #4 | 'beck anxiety inventor*':ti,ab,kw OR bai:ti,ab,kw OR 'hamilton anxiety rating':ti,ab,kw OR 'ham a':ti,ab,kw OR 'state-trait anxiety inventory':ti,ab,kw OR stai:ti,ab,kw OR 'gad 7':ti,ab,kw OR 'liebowitz social anxiety scale*':ti,ab,kw OR lsas*:ti,ab,kw OR 'penn state worry questionnaire*':ti,ab,kw OR pswq:ti,ab,kw | 38946 |
| #5 | #1 OR #2 OR #3 OR #4 | 214253 |
| #6 | 'mindfulness-based stress reduction'/de | 1857 |
| #7 | mbsr:ti,ab,kw OR 'mindfulness-based stress reduction':ti,ab,kw | 3190 |
| #8 | (mindfulness-based NEAR/3 stress ):ti,ab,kw | 3373 |
| #9 | 'mindfulness'/exp AND 'stress reduction':ti,ab,kw | 3134 |

| | | |
|---|---|---|
| #10 | #6 OR #7 OR #8 OR #9 | 4184 |
| #11 | #5 AND #10 | 554 |

| | |
|---|---|
| Database | Scopus |
| Platform | Elsevier |
| Date of last search | October 17, 2025 |
| Number of results | 323 |
| Notes | |
| #1 | ( ( TITLE-ABS-KEY ( GAD OR "Generalized Anxiet*" OR "General* Anxiety Disorder*" OR "Overanxious Disorder*" ) ) OR ( TITLE-ABS-KEY ( ( PHQ* OR "patient health questionnaire*" OR "Clinical Global Impression*" OR CGI* OR SCID OR "structured clinical interview" OR "Depression Anxiety Stress Scale*" OR DASS OR "Hospital Anxiety and Depression Scale*" OR HADS ) AND ( anxi* OR GAD ) ) ) OR ( ( TITLE-ABS-KEY ( "Beck anxiety inventor*" OR BAI OR "Hamilton Anxiety Rating" OR HAM-A OR "State-Trait Anxiety Inventory" OR STAI OR GAD-7 OR "Liebowitz Social Anxiety Scale*" OR LSAS* OR "Penn State Worry Questionnaire*" OR PSWQ ) ) ) ) AND ( ( TITLE-ABS-KEY ( MBSR OR "mindfulness-based stress reduction" OR MBSR / MBCT ) ) OR ( TITLE-ABS-KEY ( mindfulness-based W/3 stress ) ) OR ( INDEXTERMS ( mindfulness ) AND TITLE-ABS-KEY ( "stress reduction" ) ) ) |

| | | |
|---|---|---|
| Database | CINAHL | |
| Platform | EBSCO | |
| Date of last search | October 22, 2025 | |
| Number of results | 74 | |
| Notes | | |
| S1 | (MH "Anxiety Disorders") OR (MH "Generalized Anxiety Disorder") | 13567 |
| S2 | ((TI GAD OR AB GAD OR SU GAD) OR (TI "Generalized Anxiet*" OR AB "Generalized Anxiet*" OR SU "Generalized Anxiet*") OR (TI "General* Anxiety Disorder*" OR AB "General* Anxiety Disorder*" OR SU "General* Anxiety Disorder*") OR (TI "Overanxious | 6242 |

| | | |
|---|---|---|
| | Disorder*" OR AB "Overanxious Disorder*" OR SU "Overanxious Disorder*")) | |
| S3 | (((TI PHQ* OR AB PHQ* OR SU PHQ*) OR (TI "patient health questionnaire*" OR AB "patient health questionnaire*" OR SU "patient health questionnaire*") OR (TI "Clinical Global Impression*" OR AB "Clinical Global Impression*" OR SU "Clinical Global Impression*") OR (TI CGI* OR AB CGI* OR SU CGI*) OR (TI SCID OR AB SCID OR SU SCID) OR (TI "structured clinical interview" OR AB "structured clinical interview" OR SU "structured clinical interview") OR (TI "Depression Anxiety Stress Scale*" OR AB "Depression Anxiety Stress Scale*" OR SU "Depression Anxiety Stress Scale*") OR (TI DASS OR AB DASS OR SU DASS) OR (TI "Hospital Anxiety and Depression Scale*" OR AB "Hospital Anxiety and Depression Scale*" OR SU "Hospital Anxiety and Depression Scale*") OR (TI HADS OR AB HADS OR SU HADS)) AND ((TI anxi* OR AB anxi* OR SU anxi*) OR (TI GAD OR AB GAD OR SU GAD))) | 38522 |
| S4 | ((TI "Beck anxiety inventor*" OR AB "Beck anxiety inventor*" OR SU "Beck anxiety inventor*") OR (TI BAI OR AB BAI OR SU BAI) OR (TI "Hamilton Anxiety Rating" OR AB "Hamilton Anxiety Rating" OR SU "Hamilton Anxiety Rating") OR (TI HAM-A OR AB HAM-A OR SU HAM-A) OR (TI "State-Trait Anxiety Inventory" OR AB "State-Trait Anxiety Inventory" OR SU "State-Trait Anxiety Inventory") OR (TI STAI OR AB STAI OR SU STAI) OR (TI GAD-7 OR AB GAD-7 OR SU GAD-7) OR (TI "Liebowitz Social Anxiety Scale*" OR AB "Liebowitz Social Anxiety Scale*" OR SU "Liebowitz Social Anxiety Scale*") OR (TI LSAS* OR AB LSAS* OR SU LSAS*) OR (TI "Penn State Worry Questionnaire*" OR AB "Penn State Worry Questionnaire*" OR SU "Penn State Worry Questionnaire*") OR (TI PSWQ OR AB PSWQ OR SU PSWQ)) | 11531 |
| S5 | S4 OR S3 OR S2 OR S1 | 57944 |
| S6 | ((TI MBSR OR AB MBSR OR SU MBSR) OR (TI "mindfulness-based stress reduction" OR AB "mindfulness-based stress reduction" OR SU "mindfulness-based stress | 1066 |

| | | |
|---|---|---|
| | reduction") OR (TI MBSR/MBCT OR AB MBSR/MBCT OR SU MBSR/MBCT)) | |
| S7 | ((TI mindfulness-based OR AB mindfulness-based OR SU mindfulness-based) N3 (TI stress OR AB stress OR SU stress)) | 1119 |
| S8 | (MH mindfulness) AND (TI "stress reduction" OR AB "stress reduction" OR SU "stress reduction") | 800 |
| S9 | S6 OR S7 OR S8 | 1286 |
| S10 | S5 AND S9 | 74 |

| | | |
|---|---|---|
| Database | ERIC | |
| Platform | EBSCO | |
| Date of last search | 10/23/2025 | |
| Number of results | 11 | |
| Notes | | |
| S1 | DE "Anxiety Disorders" | 1159 |
| S2 | ((TI GAD OR AB GAD OR SU GAD) OR (TI "Generalized Anxiet*" OR AB "Generalized Anxiet*" OR SU "Generalized Anxiet*") OR (TI "General* Anxiety Disorder*" OR AB "General* Anxiety Disorder*" OR SU "General* Anxiety Disorder*") OR (TI "Overanxious Disorder*" OR AB "Overanxious Disorder*" OR SU "Overanxious Disorder*")) | 415 |
| S3 | (((TI PHQ* OR AB PHQ* OR SU PHQ*) OR (TI "patient health questionnaire*" OR AB "patient health questionnaire*" OR SU "patient health questionnaire*") OR (TI "Clinical Global Impression*" OR AB "Clinical Global Impression*" OR SU "Clinical Global Impression*") OR (TI CGI* OR AB CGI* OR SU CGI*) OR (TI SCID OR AB SCID OR SU SCID) OR (TI "structured clinical interview" OR AB "structured clinical interview" OR SU "structured clinical interview") OR (TI "Depression Anxiety Stress Scale*" OR AB "Depression Anxiety Stress Scale*" OR SU "Depression Anxiety Stress Scale*") OR (TI DASS OR AB DASS OR SU DASS) OR (TI "Hospital Anxiety and Depression Scale*" OR AB "Hospital Anxiety and Depression Scale*" OR SU "Hospital Anxiety and Depression Scale*") OR (TI HADS OR AB HADS OR SU | 4377 |

| | | |
|---|---|---|
| | HADS)) AND ((TI anxi* OR AB anxi* OR SU anxi*) OR (TI GAD OR AB GAD OR SU GAD))) | |
| S4 | ((TI "Beck anxiety inventor*" OR AB "Beck anxiety inventor*" OR SU "Beck anxiety inventor*") OR (TI BAI OR AB BAI OR SU BAI) OR (TI "Hamilton Anxiety Rating" OR AB "Hamilton Anxiety Rating" OR SU "Hamilton Anxiety Rating") OR (TI HAM-A OR AB HAM-A OR SU HAM-A) OR (TI "State-Trait Anxiety Inventory" OR AB "State-Trait Anxiety Inventory" OR SU "State-Trait Anxiety Inventory") OR (TI STAI OR AB STAI OR SU STAI) OR (TI GAD-7 OR AB GAD-7 OR SU GAD-7) OR (TI "Liebowitz Social Anxiety Scale*" OR AB "Liebowitz Social Anxiety Scale*" OR SU "Liebowitz Social Anxiety Scale*") OR (TI LSAS* OR AB LSAS* OR SU LSAS*) OR (TI "Penn State Worry Questionnaire*" OR AB "Penn State Worry Questionnaire*" OR SU "Penn State Worry Questionnaire*") OR (TI PSWQ OR AB PSWQ OR SU PSWQ)) | 1020 |
| S5 | S4 OR S3 OR S2 OR S1 | 6348 |
| S6 | ((TI MBSR OR AB MBSR OR SU MBSR) OR (TI "mindfulness-based stress reduction" OR AB "mindfulness-based stress reduction" OR SU "mindfulness-based stress reduction") OR (TI MBSR/MBCT OR AB MBSR/MBCT OR SU MBSR/MBCT)) | 74 |
| S7 | ((TI mindfulness-based OR AB mindfulness-based OR SU mindfulness-based) N3 (TI stress OR AB stress OR SU stress)) | 83 |
| S8 | (DE "mindfulness") AND (TI "stress reduction" OR AB "stress reduction" OR SU "stress reduction") | 0 |
| S9 | S6 OR S7 OR S8 | 86 |
| S10 | S5 AND S9 | 11 |

| | | |
|---|---|---|
| Database | SOCINDEX | |
| Platform | EBSCO | |
| Date of last search | October 23, 2025 | |
| Number of results | 12 | |
| Notes | | |
| S1 | DE "Anxiety Disorders" OR DE "Generalized Anxiety Disorder" | 2424 |
| S2 | ((TI GAD OR AB GAD OR SU GAD) OR (TI "Generalized Anxiet*" OR AB "Generalized Anxiet*" OR SU "Generalized Anxiet*") OR (TI "General* Anxiety Disorder*" OR AB "General* Anxiety Disorder*" OR SU "General* Anxiety Disorder*") OR (TI "Overanxious Disorder*" OR AB "Overanxious Disorder*" OR SU "Overanxious Disorder*")) | 1346 |
| S3 | (((TI PHQ* OR AB PHQ* OR SU PHQ*) OR (TI "patient health questionnaire*" OR AB "patient health questionnaire*" OR SU "patient health questionnaire*") OR (TI "Clinical Global Impression*" OR AB "Clinical Global Impression*" OR SU "Clinical Global Impression*") OR (TI CGI* OR AB CGI* OR SU CGI*) OR (TI SCID OR AB SCID OR SU SCID) OR (TI "structured clinical interview" OR AB "structured clinical interview" OR SU "structured clinical interview") OR (TI "Depression Anxiety Stress Scale*" OR AB "Depression Anxiety Stress Scale*" OR SU "Depression Anxiety Stress Scale*") OR (TI DASS OR AB DASS OR SU DASS) OR (TI "Hospital Anxiety and Depression Scale*" OR AB "Hospital Anxiety and Depression Scale*" OR SU "Hospital Anxiety and Depression Scale*") OR (TI HADS OR AB HADS OR SU HADS)) AND ((TI anxi* OR AB anxi* OR SU anxi*) OR (TI GAD OR AB GAD OR SU GAD))) | 7628 |
| S4 | ((TI "Beck anxiety inventor*" OR AB "Beck anxiety inventor*" OR SU "Beck anxiety inventor*") OR (TI BAI OR AB BAI OR SU BAI) OR (TI "Hamilton Anxiety Rating" OR AB "Hamilton Anxiety Rating" OR SU "Hamilton Anxiety Rating") OR (TI HAM-A OR AB HAM-A OR SU HAM-A) OR (TI "State-Trait Anxiety Inventory" OR AB "State-Trait Anxiety Inventory" OR SU "State-Trait Anxiety Inventory") OR (TI STAI OR AB STAI OR SU | 1666 |

| | | |
|---|---|---|
| | STAI) OR (TI GAD-7 OR AB GAD-7 OR SU GAD-7) OR (TI "Liebowitz Social Anxiety Scale*" OR AB "Liebowitz Social Anxiety Scale*" OR SU "Liebowitz Social Anxiety Scale*") OR (TI LSAS* OR AB LSAS* OR SU LSAS*) OR (TI "Penn State Worry Questionnaire*" OR AB "Penn State Worry Questionnaire*" OR SU "Penn State Worry Questionnaire*") OR (TI PSWQ OR AB PSWQ OR SU PSWQ)) | |
| S5 | S4 OR S3 OR S2 OR S1 | 11351 |
| S6 | ((TI MBSR OR AB MBSR OR SU MBSR) OR (TI "mindfulness-based stress reduction" OR AB "mindfulness-based stress reduction" OR SU "mindfulness-based stress reduction") OR (TI MBSR/MBCT OR AB MBSR/MBCT OR SU MBSR/MBCT)) | 135 |
| S7 | ((TI mindfulness-based OR AB mindfulness-based OR SU mindfulness-based) N3 (TI stress OR AB stress OR SU stress)) | 135 |
| S8 | (MH mindfulness) AND (TI "stress reduction" OR AB "stress reduction" OR SU "stress reduction") | 113 |
| S9 | S6 OR S7 OR S8 | 170 |
| S10 | S5 AND S9 | 12 |

| | | |
|---|---|---|
| Database | APA PsycInfo | |
| Platform | EBSCO | |
| Date of last search | October 23, 2025 | |
| Number of results | 217 | |
| Notes | | |
| S1 | DE "Anxiety Disorders" OR DE "Generalized Anxiety Disorder" | 41268 |
| S2 | ((TI GAD OR AB GAD OR SU GAD) OR (TI "Generalized Anxiet*" OR AB "Generalized Anxiet*" OR SU "Generalized Anxiet*") OR (TI "General* Anxiety Disorder*" OR AB "General* Anxiety Disorder*" OR SU "General* Anxiety Disorder*") OR (TI "Overanxious Disorder*" OR AB "Overanxious Disorder*" OR SU "Overanxious Disorder*")) | 14351 |
| S3 | (((TI PHQ* OR AB PHQ* OR SU PHQ*) OR (TI "patient health questionnaire*" OR AB "patient health | 66529 |

| | | |
|---|---|---|
| | questionnaire*" OR SU "patient health questionnaire*") OR (TI "Clinical Global Impression*" OR AB "Clinical Global Impression*" OR SU "Clinical Global Impression*") OR (TI CGI* OR AB CGI* OR SU CGI*) OR (TI SCID OR AB SCID OR SU SCID) OR (TI "structured clinical interview" OR AB "structured clinical interview" OR SU "structured clinical interview") OR (TI "Depression Anxiety Stress Scale*" OR AB "Depression Anxiety Stress Scale*" OR SU "Depression Anxiety Stress Scale*") OR (TI DASS OR AB DASS OR SU DASS) OR (TI "Hospital Anxiety and Depression Scale*" OR AB "Hospital Anxiety and Depression Scale*" OR SU "Hospital Anxiety and Depression Scale*") OR (TI HADS OR AB HADS OR SU HADS)) AND ((TI anxi* OR AB anxi* OR SU anxi*) OR (TI GAD OR AB GAD OR SU GAD))) | |
| S4 | ((TI "Beck anxiety inventor*" OR AB "Beck anxiety inventor*" OR SU "Beck anxiety inventor*") OR (TI BAI OR AB BAI OR SU BAI) OR (TI "Hamilton Anxiety Rating" OR AB "Hamilton Anxiety Rating" OR SU "Hamilton Anxiety Rating") OR (TI HAM-A OR AB HAM-A OR SU HAM-A) OR (TI "State-Trait Anxiety Inventory" OR AB "State-Trait Anxiety Inventory" OR SU "State-Trait Anxiety Inventory") OR (TI STAI OR AB STAI OR SU STAI) OR (TI GAD-7 OR AB GAD-7 OR SU GAD-7) OR (TI "Liebowitz Social Anxiety Scale*" OR AB "Liebowitz Social Anxiety Scale*" OR SU "Liebowitz Social Anxiety Scale*") OR (TI LSAS* OR AB LSAS* OR SU LSAS*) OR (TI "Penn State Worry Questionnaire*" OR AB "Penn State Worry Questionnaire*" OR SU "Penn State Worry Questionnaire*") OR (TI PSWQ OR AB PSWQ OR SU PSWQ)) | 17196 |
| S5 | S4 OR S3 OR S2 OR S1 | 111405 |
| S6 | ((TI MBSR OR AB MBSR OR SU MBSR) OR (TI "mindfulness-based stress reduction" OR AB "mindfulness-based stress reduction" OR SU "mindfulness-based stress reduction") OR (TI MBSR/MBCT OR AB MBSR/MBCT OR SU MBSR/MBCT)) | 1696 |
| S7 | ((TI mindfulness-based OR AB mindfulness-based OR SU mindfulness-based) N3 (TI stress OR AB stress OR SU stress)) | 1741 |

| | | |
|---|---|---|
| S8 | (MH mindfulness) AND (TI "stress reduction" OR AB "stress reduction" OR SU "stress reduction") | 1217 |
| S9 | S6 OR S7 OR S8 | 2034 |
| S10 | S5 AND S9 | 217 |

| | | |
|---|---|---|
| Database | Dissertations & Theses Global | |
| Platform | Proquest | |
| Date of last search | October 23, 2025 | |
| Number of results | 164 | |
| Notes | | |
| | ((TI,AB,IF(GAD) OR TI,AB,IF("Generalized Anxiet*") OR TI,AB,IF("General* Anxiety Disorder*") OR TI,AB,IF("Overanxious Disorder*")) OR ((TI,AB,IF(PHQ*) OR TI,AB,IF("patient health questionnaire*") OR TI,AB,IF("Clinical Global Impression*") OR TI,AB,IF(CGI*) OR TI,AB,IF(SCID) OR TI,AB,IF("structured clinical interview") OR TI,AB,IF("Depression Anxiety Stress Scale*") OR TI,AB,IF(DASS) OR TI,AB,IF("Hospital Anxiety and Depression Scale*") OR TI,AB,IF(HADS)) AND (TI,AB,IF(anxi*) OR TI,AB,IF(GAD))) OR ("Beck anxiety inventor*" OR BAI OR "Hamilton Anxiety Rating" OR HAM-A OR "State-Trait Anxiety Inventory" OR STAI OR GAD-7 OR "Liebowitz Social Anxiety Scale*" OR LSAS* OR "Penn State Worry Questionnaire*" OR PSWQ)) AND ((TI,AB,IF(MBSR) OR TI,AB,IF("mindfulness-based stress reduction") OR TI,AB,IF(MBSR/MBCT)) OR (TI,AB,IF(mindfulness-based) NEAR/3 TI,AB,IF(stress))) | 164 |

### Appendix 3: List of excluded trials and reasons

| Reference | Reason for exclusion |
|---|---|
| Abbasi, M., Ghorbani, N., Imani, A.H. and Tahbaz Hoseinzadeh, S. (2021), Exploring the mediating role of integrative self-knowledge in the relationship between mindfulness and well-being in the context of a mindfulness-based stress reduction program. Int J Psychol, 56: 249-256. https://doi.org/10.1002/ijop.12705 | Insufficient data to estimate GAD prevalence |
| Abd El-Maksoud M, Ahmed H, Al-Metyazidy H, Elsayed E, Aseeri A, Asiri N, Asiri M, Benjamin L, Elsalous S. Effect of Mindfulness Training Program on Promoting Mental Well-being Among Emergency Nurses . Open Nurs J, 2024; 18: e18744346327799. http://dx.doi.org/10.2174/0118744346327799240828101905 | Modifications to MBSR exceeded those specified in the preregistered protocol. |
| Acosta, Y. R. (2016). *A feasibility and acceptability study of the Spanish version of mindfulness based stress reduction* (Publication No. 10193714) [Doctoral dissertation, Alliant International University]. ProQuest Dissertations and Theses Global. | Insufficient data to estimate GAD prevalence |
| AHC Media. (2018). Mindfulness-based intervention in patients with generalized anxiety disorder. *Integrative Medicine Alert, 21*(6). | Anxiety not separately reported |
| Albright, M. G. (2023). *Effects of mindfulness-based intervention on mental health outcomes in youth who are underserved* [Doctoral dissertation, The University of Alabama at Birmingham]. | Pediatric population |
| Ali-BeigiBeni, E., Salehi, A., & Jafarry, F. (2017). The effect of mindfulness-based stress reduction program on emotional well-being (stress, anxiety, depression, and sadness) of patients with psoriasis. *Journal of Isfahan Medical School, 35*(436), 756–762. | Modifications to MBSR exceeded those specified in the preregistered protocol. |
| Alimehdi, M., Ehteshamzadeh, P., Naderi, F., Eftekharsaadi, Z., & Pasha, R. (2016). The effectiveness of Mindfulness-Based Stress Reduction on Intolerance of Uncertainty and Anxiety Sensitivity among Individuals with Generalized Anxiety Disorder. *Asian Social Science*, *12*(4), 179. https://doi.org/10.5539/ass.v12n4p179 | Modifications to MBSR exceeded those specified in the preregistered protocol. |
| Alsaqer, K., Al-Maghaireh, D., Kawafha, M. et al. The Effects of Mindfulness-Based Stress Reduction on Depression, Anxiety, Stress, and Mindfulness Among Undergraduate Nursing Students in the Psychiatric Clinical Practice. Mindfulness 16, 1250–1257 (2025). https://doi.org/10.1007/s12671-025-02559-y | Insufficient data to estimate GAD prevalence |
| Alsubaie, M., Dickens, C., Dunn, B.D. et al. Feasibility and Acceptability of Mindfulness-based Cognitive Therapy | Low prevalence of generalized anxiety |

| Reference | Reason for exclusion |
|---|---|
| Compared with Mindfulness-based Stress Reduction and Treatment as Usual in People with Depression and Cardiovascular Disorders: a Three-Arm Randomised Controlled Trial. Mindfulness 11, 30–50 (2020). | |
| An, A., Hoang, H., Trang, L., Vo, Q., Tran, L., Le, T.,…Ha, H. (2022). Investigating the effect of Mindfulness-Based Stress Reduction on stress level and brain activity of college students. *IBRO Neuroscience Reports*, *12*, 399-410. https://doi.org/10.1016/j.ibneur.2022.05.004 | Modifications to MBSR exceeded those specified in the preregistered protocol. |
| Arana, D. (2006). *The practice of mindfulness meditation to alleviate the symptoms of chronic shyness and social anxiety* (Publication No. 3221757) [Doctoral dissertation]. ProQuest Dissertations and Theses Global. | Insufficient data to estimate GAD prevalence |
| Arani, A.M., Hosseini, S., Lotfinia, S. et al. The Comparative Efficacy of Unified Transdiagnostic Protocol (UP) and Mindfulness-Based Stress Reduction Protocol (MBSR) on Emotion Regulation and Uncertainty Intolerance in Infertile Women Receiving IVF. J Clin Psychol Med Settings 30, 578–588 (2023). https://doi.org/10.1007/s10880-022-09917-1 | Did not measure anxiety |
| Arch, J. J., & Ayers, C. R. (2013). Which treatment worked better for whom? Moderators of group cognitive behavioral therapy versus adapted mindfulness based stress reduction for anxiety disorders. *Behaviour Research and Therapy*, *51*(8), 434-442. https://doi.org/https://doi.org/10.1016/j.brat.2013.04.004 | Modifications to MBSR exceeded those specified in the preregistered protocol. |
| Arch, J. J., Ayers, C. R., Baker, A., Almklov, E., Dean, D. J., & Craske, M. G. (2013). Randomized clinical trial of adapted mindfulness-based stress reduction versus group cognitive behavioral therapy for heterogeneous anxiety disorders. *Behaviour Research and Therapy*, *51*(4), 185-196. https://doi.org/https://doi.org/10.1016/j.brat.2013.01.003 | Modifications to MBSR exceeded those specified in the preregistered protocol. |
| Armani Kian, A., Vahdani, B., Noorbala, A. A., Nejatisafa, A., Arbabi, M., Zenoozian, S., Nakhjavani, M., The Impact of Mindfulness-Based Stress Reduction on Emotional Wellbeing and Glycemic Control of Patients with Type 2 Diabetes Mellitus, Journal of Diabetes Research, 2018, 1986820, 6 pages, 2018. https://doi.org/10.1155/2018/1986820 | Insufficient data to estimate GAD prevalence |
| Armstrong, B. F., III. (2019). *An affect in its own right: Investigating the case for the independence of social safeness from PA and NA* [Doctoral dissertation, McGill University]. | Low prevalence of generalized anxiety |
| Aydin, Ş., & Budak, F. K. (2023). Effect of Mindfulness-Based Stress Reduction Training on Anxiety, Depression, and Hopelessness in Menopausal Women: An Experimental Study. *Psychiatric Annals*, *53*(2), 83-94. https://doi.org/doi:10.3928/00485713-20230207-06 | Modifications to MBSR exceeded those specified in the preregistered protocol. |

| Reference | Reason for exclusion |
| --- | --- |
| Babak A, Motamedi N, Mousavi SZ, Ghasemi Darestani N. Effects of Mindfulness-Based Stress Reduction on Blood Pressure, Mental Health, and Quality of Life in Hypertensive Adult Women: A Randomized Clinical Trial Study. J Teh Univ Heart Ctr 2022;17(3):127-133. | Insufficient data to estimate GAD prevalence |
| Babak, A., Motamedi, N., Mousavi, S. Z., & Ghasemi Darestani, N. (2022). Effects of mindfulness-based stress reduction on blood pressure, mental health, and quality of life in hypertensive adult women: A randomized clinical trial study. *The Journal of Tehran Heart Center, 17*(3), 127–133. https://doi.org/10.18502/jthc.v17i3.10845 | Insufficient data to estimate GAD prevalence |
| Bennett, K., Dorjee, D. The Impact of a Mindfulness-Based Stress Reduction Course (MBSR) on Well-Being and Academic Attainment of Sixth-form Students. Mindfulness 7, 105–114 (2016). https://doi.org/10.1007/s12671-015-0430-7 | Pediatric population |
| Biegel, G. M., Brown, K. W., Shapiro, S. L., & Schubert, C. M. (2009). Mindfulness-based stress reduction for the treatment of adolescent psychiatric outpatients: A randomized clinical trial. Journal of Consulting and Clinical Psychology, 77(5), 855–866. https://doi.org/10.1037/a0016241 | Pediatric population |
| Birtwell, K., Dubrow-Marshall, L., Dubrow-Marshall, R., Duerden, T., & Dunn, A. (2017). A mixed methods evaluation of a Mindfulness-Based Stress Reduction course for people with Parkinson's disease. *Complementary Therapies in Clinical Practice*, *29*, 220-228. https://doi.org/https://doi.org/10.1016/j.ctcp.2017.10.009 | Insufficient data to estimate GAD prevalence |
| Blard, F., Erraud, J., Bonnet, M., Auzou, N., & Tison, F. (2024). Mindfulness in Parkinson's disease: A French national survey and a pilot intervention feasibility trial using the MBSR program (M-Park). *Revue Neurologique*, *180*(8), 777-790. https://doi.org/https://doi.org/10.1016/j.neurol.2024.03.013 | Insufficient data to estimate GAD prevalence |
| Bossi, F., Zaninotto, F., D'Arcangelo, S. et al. Mindfulness-based online intervention increases well-being and decreases stress after Covid-19 lockdown. Sci Rep 12, 6483 (2022). https://doi.org/10.1038/s41598-022-10361-2 | Modifications to MBSR exceeded those specified in the preregistered protocol. |
| Bratton, B. L. (2008). *Mindfulness in childbirth: An investigation of the effects of mindfulness training on maternal satisfaction with childbirth and obstetric outcomes* [Doctoral dissertation, Western Michigan University]. ScholarWorks at WMU. https://scholarworks.wmich.edu/dissertations/754 | Modifications to MBSR exceeded those specified in the preregistered protocol. |
| Brito Pons, G. (2014). *Cultivating healthy minds and open hearts: A mixed-method controlled study on the psychological and relational effects of compassion cultivation training in Chile* (Publication No. 3631418) [Doctoral dissertation, Sofia University]. ProQuest Dissertations and Theses Global. | Insufficient data to estimate GAD prevalence |

| Reference | Reason for exclusion |
|---|---|
| Brown MM, Arigo D, Wolever RQ, et al. Do gender, anxiety, or sleep quality predict mindfulness-based stress reduction outcomes? Journal of Health Psychology. 2021;26(13):2656-2662. doi:10.1177/1359105320931186 | Anxiety not separately reported |
| Brown, K. W., West, A. M., Loverich, T. M., & Biegel, G. M. (2011). Assessing adolescent mindfulness: Validation of an Adapted Mindful Attention Awareness Scale in adolescent normative and psychiatric populations. Psychological Assessment, 23(4), 1023–1033. https://doi.org/10.1037/a0021338 | Pediatric population |
| Burrowes, S. (2018). *The effects of mindfulness based stress reduction on brain gray matter volume and psychosocial co-morbidities in episodic migraine patients* (Publication No. 10935161) [Doctoral dissertation, University of Maryland, Baltimore]. ProQuest Dissertations and Theses Global. | Modifications to MBSR exceeded those specified in the preregistered protocol. |
| Burrowes, S. A. B., Goloubeva, O., Stafford, K., McArdle, P. F., Goyal, M., Peterlin, B. L., Haythornthwaite, J. A., & Seminowicz, D. A. (2021). Enhanced mindfulness-based stress reduction in episodic migraine—effects on sleep quality, anxiety, stress, and depression: a secondary analysis of a randomized clinical trial. Pain, 163(3), 436–444. https://doi.org/10.1097/j.pain.0000000000002372 | Modifications to MBSR exceeded those specified in the preregistered protocol. |
| Canella, R., Fazia, T., & Gardi, C. (2025). Mindfulness-based stress reduction program improves psychological well-being and blood pressure in an Italian context: potential mechanisms and benefits. Psychology, Health & Medicine, 1–16. | Insufficient data to estimate GAD prevalence |
| Capobianco, L., Reeves, D., Morrison, A. P., & Wells, A. (2018). Group Metacognitive Therapy vs. Mindfulness Meditation Therapy in a Transdiagnostic Patient Sample: A Randomised Feasibility Trial. *Psychiatry Research*, *259*, 554-561. https://doi.org/https://doi.org/10.1016/j.psychres.2017.11.045 | Modifications to MBSR exceeded those specified in the preregistered protocol. |
| Cary, E. L., Bergen-Cico, D., Sinegar, S., Schutt, M. K. A., Helminen, E. C., & Felver, J. C. (2024). Self-regulation mediates effects of adapted mindfulness-based stress reduction on anxiety among college students. Journal of American College Health, 72(9), 3818–3828. https://doi.org/10.1080/07448481.2023.2201843 | Modifications to MBSR exceeded those specified in the preregistered protocol. |
| Casas, I.; Nakaki, A.; Pascal,R.; Castro-Barquero, S.; Youssef, L.;Genero, M.; Benitez, L.; Larroya, M.;Boutet, M.L.; Casu, G.; et al. Effects of a Mediterranean Diet Intervention on Maternal Stress, Well-Being, and Sleep Quality throughout Gestation—TheIMPACT-BCN Trial.Nutrients2023,15, 2362 | Intervention is not MBSR |
| Chang, Y.-C., Chiu, C.-F., Wang, C.-K., Wu, C.-T., Liu, L.-C., Wu, Y.-C., 2022. Short-term effect of internet-delivered | Insufficient data to estimate GAD prevalence |

| Reference | Reason for exclusion |
|---|---|
| mindfulness-based stress reduction on mental health, self-efficacy, and body image among women with breast cancer during the COVID-19 pandemic. Frontiers in Psychology 13. | |
| Chang, YC., Lin, GM., Yeh, TL. et al. Impact of mindfulness-based stress reduction on female sexual function and mental health in patients with breast cancer. Support Care Cancer 30, 4315–4325 (2022). https://doi.org/10.1007/s00520-021-06540-y | Insufficient data to estimate GAD prevalence |
| Chen, J. A., Anderson, M. L., Cherkin, D. C., Balderson, B. H., Cook, A. J., Sherman, K. J., & Turner, J. A. (2023). Moderators and Nonspecific Predictors of Treatment Benefits in a Randomized Trial of Mindfulness-Based Stress Reduction vs Cognitive-Behavioral Therapy vs Usual Care for Chronic Low Back Pain. *The Journal of Pain*, *24*(2), 282-303. https://doi.org/10.1016/j.jpain.2022.09.014 | Low prevalence of generalized anxiety |
| Choi, H. S. (2022). *The impact of selective attention on energy and its implications for health* [Doctoral dissertation, Columbia University]. | Intervention is not MBSR |
| Choles, J. R. (2018). *Impacts of mindfulness training on mechanisms underlying stress reduction in teachers: Results from a randomized controlled trial* (Publication No. 10750824) [Master's thesis, Portland State University]. ProQuest Dissertations and Theses Global. | Modifications to MBSR exceeded those specified in the preregistered protocol. |
| Ciacchini, R.; Conversano,C.; Orrù, G.; Scafuto, F.; Sabbatini, S.;Paroli, M.; Miniati, M.; Matiz, A.;Gemignani, A.; Crescentini, C. AboutDistress in Chronic Pain Conditions:A Pre–Post Study on the Effectivenessof a Mindfulness-Based Interventionfor Fibromyalgia and Low Back PainPatients.Int. J. Environ. Res. PublicHealth2024,21, 1507 | Insufficient data to estimate GAD prevalence |
| Collado-Navarro, C., Navarro-Gil, M., Pérez-Aranda, A., López-del-Hoyo, Y., Garcia-Campayo, J., & Montero-Marin, J. (2021). Effectiveness of mindfulness-based stress reduction and attachment-based compassion therapy for the treatment of depressive, anxious, and adjustment disorders in mental health settings: A randomized controlled trial. Depression Anxiety, 38, 1138–1151. https://doi.org/10.1002/da.23198 | Low prevalence of generalized anxiety |
| Correia, P., Quaresma, C., Morais, P., Fonseca, M., Vigário, R., & Quintão, C. (2023). Effects of mindfulness-based stress reduction meditation on the emotional reaction to affective pictures assessed by electrodermal activity. *Biomedical Signal Processing and Control*, *86*, 105314. https://doi.org/https://doi.org/10.1016/j.bspc.2023.105314 | Insufficient data to estimate GAD prevalence |
| de Morais, J. P. A. N. M. (2021). *O efeito da meditação mindfulness na actividade electrofisiológica: Avaliação do* | Insufficient data to estimate GAD prevalence |

| Reference | Reason for exclusion |
|---|---|
| *estado de concentração, controlo emocional e qualidade de vida* [The effect of mindfulness meditation on electrophysiological activity: Assessment of concentration state, emotional control, and quality of life] [Doctoral dissertation, Universidade Nova de Lisboa]. | |
| Díaz-González, M. C., Pérez Dueñas, C., Sánchez-Raya, A., Moriana Elvira, J. A., & Sánchez Vázquez, V. (2018). Mindfulness-based stress reduction in adolescents with mental disorders: A randomised clinical trial. *Psicothema, 30*(2), 165–170. https://doi.org/10.7334/psicothema2017.259 | Pediatric population |
| dos Santos, T. M., Kozasa, E. H., Carmagnani, I. S., Tanaka, L. H., Lacerda, S. S., & Nogueira-Martins, L. A. (2016). Positive Effects of a Stress Reduction Program Based on Mindfulness Meditation in Brazilian Nursing Professionals: Qualitative and Quantitative Evaluation. *EXPLORE*, *12*(2), 90-99. https://doi.org/https://doi.org/10.1016/j.explore.2015.12.005 | Modifications to MBSR exceeded those specified in the preregistered protocol. |
| Dundas, I., Thorsheim, T., Hjeltnes, A., & Binder, P. E. (2016). Mindfulness Based Stress Reduction for Academic Evaluation Anxiety: A Naturalistic Longitudinal Study. Journal of College Student Psychotherapy, 30(2), 114–131. https://doi.org/10.1080/87568225.2016.1140988 | Insufficient data to estimate GAD prevalence |
| Edwards, M., Adams, E. M., Waldo, M., Hadfield, O. D., & Biegel, G. M. (2014). Effects of a Mindfulness Group on Latino Adolescent Students: Examining Levels of Perceived Stress, Mindfulness, Self-Compassion, and Psychological Symptoms. The Journal for Specialists in Group Work, 39(2), 145–163. https://doi.org/10.1080/01933922.2014.891683 | Pediatric population |
| Elimimian E, Elson L, Bilani N, et al. Long-Term Effect of a Nonrandomized Psychosocial Mindfulness-Based Intervention in Hispanic/Latina Breast Cancer Survivors. Integrative Cancer Therapies. 2020;19. doi:10.1177/1534735419890682 | Low prevalence of generalized anxiety |
| Epstein, B. J. (2009). *Effects of a mindfulness based stress reduction program on fathers of children with developmental disability* (Publication No. 3421917) [Doctoral dissertation, Hofstra University]. ProQuest Dissertations and Theses Global. | Modifications to MBSR exceeded those specified in the preregistered protocol. |
| Fariba Eslamimoghadam, Zahra Abedini, Ashraf Khoramirad, The mental health of nursing students during the COVID-19 pandemic: Beneficial effects of mindfulness-based stress reduction, Heliyon, Volume 10, Issue 12, 2024, e32986, ISSN 2405-8440, | Modifications to MBSR exceeded those specified in the preregistered protocol. |
| Fendel JC, Aeschbach VM, Schmidt S, Göritz AS. The impact of a tailored mindfulness-based program for resident physicians on distress and the quality of care: A randomised controlled trial. J Intern Med. 2021; 290: 1233–1248. | Low prevalence of generalized anxiety |

| Reference | Reason for exclusion |
| --- | --- |
| Ferren, D. J. (2003). *The clinical use of mindfulness meditation within an intensive psychotherapy program for clients diagnosed with a personality disorder* [Doctoral dissertation, University of Toronto]. | Insufficient data to estimate GAD prevalence |
| Formeister E J, Mitchell J, Krauter R, et al. (February 18, 2025) Mindfulness-Based Stress Reduction for the Treatment of Vestibular Migraine: A Prospective Pilot Study. Cureus 17(2): e79225. | Insufficient data to estimate GAD prevalence |
| Foureur, M., Besley, K., Burton, G., Yu, N., & Crisp, J. (2013). Enhancing the resilience of nurses and midwives: Pilot of a mindfulnessbased program for increased health, sense of coherence and decreased depression, anxiety and stress. Contemporary Nurse, 45(1), 114–125. https://doi.org/10.5172/conu.2013.45.1.114 | Modifications to MBSR exceeded those specified in the preregistered protocol. |
| Freedenberg, V. A. (2013). *Feasibility and effect of a mindfulness based stress reduction program on anxiety, depression, and coping in a sample of adolescents with implantable cardioverter defibrillators or pacemakers* [Doctoral dissertation, University of Maryland, Baltimore]. | Pediatric population |
| Gallego J, Aguilar-Parra JM, Cangas AJ, Langer ÁI, Mañas I. Effect of a Mindfulness Program on Stress, Anxiety and Depression in University Students. The Spanish Journal of Psychology. 2014;17:E109. doi:10.1017/sjp.2014.102 | Modifications to MBSR exceeded those specified in the preregistered protocol. |
| Gans, J.J., O'Sullivan, P. & Bircheff, V. Mindfulness Based Tinnitus Stress Reduction Pilot Study. Mindfulness 5, 322–333 (2014). https://doi.org/10.1007/s12671-012-0184-4 | Modifications to MBSR exceeded those specified in the preregistered protocol. |
| García-Campayo, J., Andrés-Rodríguez, L., Borràs, X., Rozadilla-Sacanell, A., Peñarrubia-Maria, M. T., Angarita-Osorio, N., McCracken, L. M., Luciano, J. V., Pérez-Aranda, A., Feliu-Soler, A., & Montero-Marín, J. (2019). A randomized controlled efficacy trial of mindfulness-based stress reduction compared with an active control group and usual care for fibromyalgia: the EUDAIMON study. Pain, 160(11), 2508–2523. https://doi.org/10.1097/j.pain.0000000000001655 | Anxiety not separately reported |
| Gerrity, P., Waite, R., Metzker, K., & Smith, B. (2015). Short-Form Mindfulness-Based Stress Reduction Reduces Anxiety and Improves Health-Related Quality of Life in an Inner-City Population. Holistic Nursing Practice, 29(2), 70–77. https://doi.org/10.1097/HNP.0000000000000075 | Modifications to MBSR exceeded those specified in the preregistered protocol. |
| Goldin, P. R., & Gross, J. J. (2010). Effects of mindfulness-based stress reduction (MBSR) on emotion regulation in social anxiety disorder. Emotion, 10(1), 83–91. https://doi.org/10.1037/a0018441 | Low prevalence of generalized anxiety |
| Goldin, P. R., Morrison, A., Jazaieri, H., Brozovich, F., Heimberg, R., & Gross, J. J. (2016). Group CBT versus MBSR | Low prevalence of generalized anxiety |

| Reference | Reason for exclusion |
|---|---|
| for social anxiety disorder: A randomized controlled trial. Journal of Consulting and Clinical Psychology, 84(5), 427–437. https://doi.org/10.1037/ccp0000092 | |
| Goldin, P., Ramel, W., & Gross, J. Mindfulness Meditation Training and Self-Referential Processing in Social Anxiety Disorder: Behavioral and Neural Effects. *J Cogn Psychother*(3), 242-257. https://doi.org/10.1891/0889-8391.23.3.242 | Low prevalence of generalized anxiety |
| Goldin, P., Ziv, M., Jazaieri, H., Hahn, K., & Gross, J. J. (2013). MBSR vs aerobic exercise in social anxiety: fMRI of emotion regulation of negative self-beliefs. *Social Cognitive and Affective Neuroscience*, *8*(1), 65-72. https://doi.org/10.1093/scan/nss054 | Low prevalence of generalized anxiety |
| Gordon, J. L., Halleran, M., Beshai, S., Eisenlohr-Moul, T. A., Frederick, J., & Campbell, T. S. (2021). Endocrine and psychosocial moderators of mindfulness-based stress reduction for the prevention of perimenopausal depressive symptoms: A randomized controlled trial. *Psychoneuroendocrinology*, *130*, 105277. https://doi.org/https://doi.org/10.1016/j.psyneuen.2021.105277 | Insufficient data to estimate GAD prevalence |
| Gotink, R. A., Younge, J. O., Wery, M. F., Utens, E. M. W. J., Michels, M., Rizopoulos, D.,…Hunink, M. M. G. (2017). Online mindfulness as a promising method to improve exercise capacity in heart disease: 12-month follow-up of a randomized controlled trial. *PLOS ONE*, *12*(5), e0175923. https://doi.org/10.1371/journal.pone.0175923 | Modifications to MBSR exceeded those specified in the preregistered protocol. |
| Gross, C. R., Kreitzer, M. J., Russas, V., Treesak, C., Frazier, P. A., & Hertz, M. I. (2004). Mindfulness meditation to reduce symptoms after organ transplant: A pilot study. *Alternative Therapies in Health and Medicine, 10*(3), 58–66. | Insufficient data to estimate GAD prevalence |
| Gross, C. R., Kreitzer, M. J., Thomas, W., Reilly-Spong, M., Cramer-Bornemann, M., Nyman, J. A., Frazier, P., & Ibrahim, H. N. (2010). Mindfulness-based stress reduction for solid organ transplant recipients: A randomized controlled trial. *Alternative Therapies in Health and Medicine, 16*(5), 30–38. | Insufficient data to estimate GAD prevalence |
| Gross, C. R., Reilly-Spong, M., Park, T., Zhao, R., Gurvich, O. V., & Ibrahim, H. N. (2017). Telephone-adapted Mindfulness-based Stress Reduction (tMBSR) for patients awaiting kidney transplantation. *Contemporary Clinical Trials*, *57*, 37-43. https://doi.org/https://doi.org/10.1016/j.cct.2017.03.014 | Insufficient data to estimate GAD prevalence |
| Gu, JJ., Tong, XS., Meng, SS. et al. Effect of mindfulness-based stress reduction in patients with acute myocardial infarction after successful primary percutaneous coronary intervention: a retrospective study. BMC Cardiovasc Disord 23, 315 (2023) | Observational Study |

| Reference | Reason for exclusion |
| --- | --- |
| Güney, E., Cengizhan, S. Ö., Karataş Okyay, E., Bal, Z., & Uçar, T. (2022). Effect of the Mindfulness-Based Stress Reduction program on stress, anxiety, and childbirth fear in pregnant women diagnosed with COVID-19. *Complementary Therapies in Clinical Practice*, *47*, 101566. https://doi.org/https://doi.org/10.1016/j.ctcp.2022.101566 | Modifications to MBSR exceeded those specified in the preregistered protocol. |
| Guo Huan, Yuan Jing, Zhong Yan, Xu Qinglei, Fan Guangcui, Cai Yue'e, Cheng Yanyan. Effects of mindfulness-based stress reduction on negative emotions and quality of life in patients with complex spinal metastases undergoing arterial embolization chemotherapy [J]. Oncology Pharmacy, 2019, 9(2):344-348,352. DOI:10.3969/j.issn.2095-1264.2019.02.35. | Insufficient data to estimate GAD prevalence |
| Haghighinejad, H., Ghazipoor, H., Jafari, P. et al. Investigating the impact of modified mindfulness-based stress reduction (MBSR) program on occupational burnout and other mental health status among nonmedical staff in a hospital: a randomized controlled trial. Int Arch Occup Environ Health 95, 2005–2016 (2022). | Modifications to MBSR exceeded those specified in the preregistered protocol |
| Hankin, V. M. (2009). *Mindfulness based stress reduction in couples facing multiple sclerosis: Impact on self reported anxiety and uncertainty* (Publication No. 3378573) [Doctoral dissertation, The City University of New York]. ProQuest Dissertations and Theses Global. | Insufficient data to estimate GAD prevalence |
| Harp, N.R., Blair, R.J.R. & Neta, M. Shift in Valence Bias Associated with Decrease in Trait Anxiety and Depression Symptoms. Cogn Ther Res 48, 444–450 (2024). https://doi.org/10.1007/s10608-023-10437-x | Insufficient data to estimate GAD prevalence |
| Hassanvandi, S., Mohammadzadegan, R., Saadat, S.H. et al. Effectiveness of Mindfulness-Based Stress Reduction on Salivary Cortisol and α-amylase Level, Anxiety and Depression in Students: A Randomized and Parallel-Group Clinical Trial. Appl Psychophysiol Biofeedback 50, 417–428 (2025) | Modifications to MBSR exceeded those specified in the preregistered protocol. |
| Higgins, E. T., Davidson, R. J., Busse, W. W., Klaus, D. R., Bednarek, G. T., Goldman, R. I.,…Rosenkranz, M. A. (2022). Clinically relevant effects of Mindfulness-Based Stress Reduction in individuals with asthma. *Brain, Behavior, & Immunity - Health*, *25*, 100509. https://doi.org/https://doi.org/10.1016/j.bbih.2022.100509 | Insufficient data to estimate GAD prevalence |
| Hoffman, C. J., Ersser, S. J., Hopkinson, J. B., Nicholls, P. G., Harrington, J. E., & Thomas, P. W. (2012). Effectiveness of Mindfulness-Based Stress Reduction in Mood, Breast- and Endocrine-Related Quality of Life, and Well-Being in Stage 0 to III Breast Cancer: A Randomized, Controlled Trial. *Journal of Clinical Oncology*, *30*(12), 1335-1342. https://doi.org/10.1200/jco.2010.34.0331 | Insufficient data to estimate GAD prevalence |

| Reference | Reason for exclusion |
|---|---|
| Hoge EA, Reese HE, Oliva IA, Gabriel CD, Guidos BM, Bui E, Simon NM and Dutton MA (2020) Investigating the Role of Interpretation Bias in Mindfulness-Based Treatment of Adults With Generalized Anxiety Disorder. Front. Psychol. 11:82. doi: 10.3389/fpsyg.2020.00082 | Subset of an already included trial |
| Hoge, E. A., Bui, E., Palitz, S. A., Schwarz, N. R., Owens, M. E., Johnston, J. M.,…Simon, N. M. (2018). The effect of mindfulness meditation training on biological acute stress responses in generalized anxiety disorder. *Psychiatry Research*, *262*, 328-332. https://doi.org/https://doi.org/10.1016/j.psychres.2017.01.006 | Anxiety not separately reported |
| Hoge, E. A., Guidos, B. M., Mete, M., Bui, E., Pollack, M. H., Simon, N. M., & Dutton, M. A. (2017). Effects of mindfulness meditation on occupational functioning and health care utilization in individuals with anxiety. *Journal of Psychosomatic Research*, *95*, 7-11. https://doi.org/https://doi.org/10.1016/j.jpsychores.2017.01.011 | Did not measure anxiety |
| Hölzel, B. K., Hoge, E. A., Greve, D. N., Gard, T., Creswell, J. D., Brown, K. W.,…Lazar, S. W. (2013). Neural mechanisms of symptom improvements in generalized anxiety disorder following mindfulness training. *NeuroImage: Clinical*, *2*, 448-458. https://doi.org/https://doi.org/10.1016/j.nicl.2013.03.011 | Anxiety not separately reported |
| Hou, J. (2012). *The effects of mindfulness-based stress reduction program on depressive symptoms reduction among family caregivers in Hong Kong* (Publication No. 3538906) [Doctoral dissertation, The Chinese University of Hong Kong]. ProQuest Dissertations and Theses Global. | Insufficient data to estimate GAD prevalence |
| Hou, J. (2012). *The effects of mindfulness-based stress reduction program on depressive symptoms reduction among family caregivers in Hong Kong* (Publication No. 3538906) [Doctoral dissertation, The Chinese University of Hong Kong]. ProQuest Dissertations and Theses Global. | Insufficient data to estimate GAD prevalence |
| Hou, R. J., Wong, S. Y. S., Yip, B. H. K., Hung, A. T. F., Lo, H. H. M., Chan, P. H. S., Lo, C. S. L., Kwok, T., Tang, W. K., Mak, W. W. S., Mercer, S. W., & Helen, S. (2013). The Effects of Mindfulness-Based Stress Reduction Program on the Mental Health of Family Caregivers: A Randomized Controlled Trial. Psychotherapy and Psychosomatics, 83(1), 45–53. https://doi.org/10.1159/000353278 | Insufficient data to estimate GAD prevalence |
| Houghton, V. T. (2008). *A quantitative study of the effectiveness of mindfulness-based stress reduction treatment, using an internet-delivered self-help program, for women with generalized anxiety disorder* (Publication No. 3310709) [Doctoral dissertation, Capella University]. ProQuest Dissertations and Theses Global. | Modifications to MBSR exceeded those specified in the preregistered protocol. |

| Reference | Reason for exclusion |
| --- | --- |
| Huang D-Z, Ismail R, Yeoh KK and Ahmad A (2025) Beyond distress: a sequential quantitative investigation of MBSR through a dual-factor model of mental health in college students. Front. Psychol. 16:1588162. | Anxiety not separately reported |
| Huang, S., Wang, Z., Zheng, D., & Liu, L. (2023). Anxiety disorder in menopausal women and the intervention efficacy of mindfulness-based stress reduction. *American Journal of Translational Research, 15*(3), 2016–2024 | Modifications to MBSR exceeded those specified in the preregistered protocol. |
| Ironmonger, E. C. (2018). *Mindfulness self-help for health care professionals* (Publication No. 27795889) [Doctoral dissertation, Canterbury Christ Church University]. ProQuest Dissertations and Theses Global. | Intervention is not MBSR |
| Isabelle Gaboury, Patricia L Dobkin, Françoise Gendron, Pasquale Roberge, Marie-Claude Beaulieu, Nathalie Carrier, Pierre Dagenais, Sophie Roux, Gilles Boire, Mindfulness-based stress reduction to improve depression, pain and high patient global assessment in controlled rheumatoid arthritis, Rheumatology Advances in Practice, Volume 6, Issue 3, 2022, rkac074 | Low prevalence of generalized anxiety |
| Iyengar, S., Sharma, M., & Parswani, M. (2013). Mindfulness-based stress reduction program in coronary heart disease: A randomized control trial. International Journal of Yoga, 6(2), 111–117. https://doi.org/10.4103/0973-6131.113405 | Modifications to MBSR exceeded those specified in the preregistered protocol. |
| Johns, S.A., Brown, L.F., Beck-Coon, K. et al. Randomized controlled pilot trial of mindfulness-based stress reduction compared to psychoeducational support for persistently fatigued breast and colorectal cancer survivors. Support Care Cancer 24, 4085–4096 (2016). https://doi.org/10.1007/s00520-016-3220-4 | Low prevalence of generalized anxiety |
| Joo, H. M., Lee, S. J., Chung, Y. G., & Shin, I. Y. (2010). Effects of Mindfulness Based Stress Reduction Program on Depression, Anxiety and Stress in Patients with Aneurysmal Subarachnoid Hemorrhage. *J Korean Neurosurg Soc*, *47*(5), 345-351. https://doi.org/10.3340/jkns.2010.47.5.345 | Insufficient data to estimate GAD prevalence |
| Juul, L., Pallesen, K.J., Bjerggaard, M. et al. A pilot randomised trial comparing a mindfulness-based stress reduction course, a locally-developed stress reduction intervention and a waiting list control group in a real-life municipal health care setting. BMC Public Health 20, 409 (2020). https://doi.org/10.1186/s12889-020-08470-6 | Anxiety not separately reported |
| Kabat-Zinn, J., Massion, A. O., Kristeller, J., Peterson, L. G., Fletcher, K. E., Pbert, L., Lenderking, W. R., & Santorelli, S. F. (1992). Effectiveness of a meditation-based stress reduction | Low prevalence of generalized anxiety |

| Reference | Reason for exclusion |
|---|---|
| program in the treatment of anxiety disorders. *American Journal of Psychiatry, 149*(7), 936–943. | |
| Keng, S.-L., Looi, P. S., Tan, E. L. Y., Yim, O.-S., Lai, P. S., Chew, S. H., & Ebstein, R. P. (2020). Effects of Mindfulness-Based Stress Reduction on Psychological Symptoms and Telomere Length: A Randomized Active-Controlled Trial. *Behavior Therapy*, *51*(6), 984-996. https://doi.org/https://doi.org/10.1016/j.beth.2020.01.005 | Low prevalence of generalized anxiety |
| Kenne Sarenmalm, E., Mårtensson, L. B., Andersson, B. A., Karlsson, P., & Bergh, I. (2017). Mindfulness and its efficacy for psychological and biological responses in women with breast cancer. *Cancer Medicine*, *6*(5), 1108-1122. https://doi.org/https://doi.org/10.1002/cam4.1052 | Low prevalence of generalized anxiety |
| Khaled Younis Albahadly, W., Mohammed, A. H. ., Sabti, A. A. ., Al-Hili, A. ., Adnan, M. ., & Sabri, S. K. . (2022). The Effectiveness of Mindfulness-Based Stress Reduction on Stress, Anxiety, and Depression of Patients with Breast Cancer. International Journal of Body, Mind and Culture, 10(1), 109–119. https://doi.org/10.22122/ijbmc.v10i1.426 | Insufficient data to estimate GAD prevalence |
| Khoramirad A, Abedini Z, Gaeeni M, Dehghani H, Akhoundzadeh K (2024), "Mindfulness-based stress reduction via mobile phone on female family caregivers' anxiety and sleep quality during COVID-19". The Journal of Mental Health Training, Education and Practice, Vol. 19 No. 6 pp. 315–324, doi: https://doi.org/10.1108/JMHTEP-05-2023-0046 | Modifications to MBSR exceeded those specified in the preregistered protocol. |
| Khoshbin, A., Tahery, N., Nekoueian, P. et al. The effect of mindfulness group counseling on the anxiety of pregnant women exposed to domestic violence: a randomized clinical trial. BMC Psychol 13, 1076 (2025) | Modifications to MBSR exceeded those specified in the preregistered protocol. |
| Klevnick, L. (2008). *An exploration of the relationship between mindfulness and forgiveness* [Doctoral dissertation, University of Toronto]. | Insufficient data to estimate GAD prevalence |
| Kolahkaj, B., & Zargar, F. (2015). Effect of mindfulness-based stress reduction on anxiety, depression and stress in women with multiple sclerosis. *Nursing and Midwifery Studies, 4*(4), e29655. https://doi.org/10.17795/nmsjournal29655 | Insufficient data to estimate GAD prevalence |
| Kornfield, S. L., Nicol, G. E., Lenze, E. J., Yingling, M., & Loebach Wetherell, J. (2022). Mindfulness-based stress reduction in older individuals: Who benefits the most? *Advances in Mind-Body Medicine, 36*(1), 4–12. | Insufficient data to estimate GAD prevalence |
| Kristofersson, G. K. (2012). *The effects of a mindfulness based intervention on impulsivity, symptoms of depression, anxiety, experiences and quality of life of persons suffering from* | Modifications to MBSR exceeded those specified in the preregistered protocol. |

| Reference | Reason for exclusion |
| --- | --- |
| *substance use disorders and traumatic brain injury* (Publication No. 3540920) [Doctoral dissertation, University of Minnesota]. ProQuest Dissertations and Theses Global. | |
| Lan, L., Liu, W., Liu, C., Wang, H., Wang, M., Abbey, C.,…Tong, L. (2024). Effects of Mindfulness and Life-Skills Training on Emotion Regulation and Anxiety Symptoms in Chinese Migrant Children: A Randomized Controlled Trial. *Journal of Adolescent Health*, *75*(3), 404-415. https://doi.org/10.1016/j.jadohealth.2024.05.024 | Pediatric population |
| Lawrence, M., Davis, B., Clark, N.E. et al. A Pilot Randomized Controlled Trial Testing the Feasibility and Acceptability of Helping Ease Anxiety and Depression after Stroke (HEADS: UP): An Online Mindfulness-Based Intervention for Stroke Survivors. Mindfulness 16, 1862–1875 (2025). https://doi.org/10.1007/s12671-025-02595-8 | Modifications to MBSR exceeded those specified in the preregistered protocol. |
| Lawrence, M., Davis, B., Clark, N.E. et al. In-person and online mixed method non-randomised studies exploring feasibility and acceptability of HEADS: UP, an adapted Mindfulness-Based Stress Reduction programme for stroke survivors experiencing symptoms of anxiety and depression. Pilot Feasibility Stud 10, 119 (2024) | Modifications to MBSR exceeded those specified in the preregistered protocol. |
| Lengacher CA, Reich RR, Kip KE, et al. Influence of Mindfulness-Based Stress Reduction (MBSR) on Telomerase Activity in Women With Breast Cancer (BC). Biological Research For Nursing. 2014;16(4):438-447. doi:10.1177/1099800413519495 | Modifications to MBSR exceeded those specified in the preregistered protocol. |
| Lengacher CA, Reich RR, Ramesar S, et al. Feasibility of the mobile mindfulness-based stress reduction for breast cancer (mMBSR(BC)) program for symptom improvement among breast cancer survivors. Psycho-Oncology. 2018; 27: 524–531. https://doi.org/10.1002/pon.4491 | Modifications to MBSR exceeded those specified in the preregistered protocol. |
| Lengacher, C.A., Johnson-Mallard, V., Post-White, J., Moscoso, M.S., Jacobsen, P.B., Klein, T.W., Widen, R.H., Fitzgerald, S.G., Shelton, M.M., Barta, M., Goodman, M., Cox, C.E. and Kip, K.E. (2009), Randomized controlled trial of mindfulness-based stress reduction (MBSR) for survivors of breast cancer. Psycho-Oncology, 18: 1261-1272. https://doi.org/10.1002/pon.1529 | Modifications to MBSR exceeded those specified in the preregistered protocol. |
| Lengacher, C.A., Shelton, M.M., Reich, R.R. et al. Mindfulness based stress reduction (MBSR(BC)) in breast cancer: evaluating fear of recurrence (FOR) as a mediator of psychological and physical symptoms in a randomized control trial (RCT). J Behav Med 37, 185–195 (2014). https://doi.org/10.1007/s10865-012-9473-6 | Modifications to MBSR exceeded those specified in the preregistered protocol. |

| Reference | Reason for exclusion |
| --- | --- |
| Lenze E. J., Hickman S., Hershey T., Wendleton L., Ly K., Dixon D., Doré P. and Wetherell J. L. (2014), Mindfulness-based stress reduction for older adults with worry symptoms and co-occurring cognitive dysfunction, International Journal of Geriatric Psychiatry, 29, pages 991–1000, doi: 10.1002/gps.4086 | Low prevalence of generalized anxiety |
| León, C., Mirapeix, R., Blasco, T., Jovell, E., Arcusa, À., Martín, A., & Seguí, M. À. (2014). Mindfulness para la reducción del malestar emocional en pacientes oncológicos. Estudio comparativo con una intervención psicoeducativa estándar. *Psicooncología*, *10*(2-3). https://doi.org/10.5209/rev_psic.2013.v10.n2-3.43448 | Insufficient data to estimate GAD prevalence |
| Lestage, P., & Xu, R. (2016). Effets de la pratique de la pleine conscience et du Tai Chi Chuan sur la santé mentale d'étudiants : une étude pilote contrôlée non randomisée. *Journal de Thérapie Comportementale et Cognitive*, *26*(1), 32-48. https://doi.org/https://doi.org/10.1016/j.jtcc.2015.12.003 | Modifications to MBSR exceeded those specified in the preregistered protocol. |
| López-del-Hoyo, Y., Collado-Navarro, C., Pérez-Aranda, A., García-Campayo, J., López-Montoyo, A., Feliu-Soler, A.,…Montero-Marin, J. (2022). Assessing mindfulness and self-compassion facets as mediators of change in patients with depressive, anxious and adjustment disorders: Secondary data analysis of a randomized controlled trial. *Journal of Contextual Behavioral Science*, *24*, 171-178. https://doi.org/https://doi.org/10.1016/j.jcbs.2022.05.007 | Insufficient data to estimate GAD prevalence |
| Lundgren, O. (2018). *Psychological Resources and Risk Factors in Coronary Heart Disease : Assessment, Impact and the Influence of Mindfulness Training* (Publication Number 1623) [Doctoral thesis, comprehensive summary, Linköping University Electronic Press]. DiVA. Linköping. https://doi.org/10.3384/diss.diva-148332http://urn.kb.se/resolve?urn=urn:nbn:se:liu:diva-148332 | Low prevalence of generalized anxiety |
| Lush, E., Salmon, P., Floyd, A. et al. Mindfulness Meditation for Symptom Reduction in Fibromyalgia: Psychophysiological Correlates. J Clin Psychol Med Settings 16, 200–207 (2009). https://doi.org/10.1007/s10880-009-9153-z | Insufficient data to estimate GAD prevalence |
| Marais, C., Song, Y., Ferreira, R. et al. Evaluation of mindfulness based stress reduction in symptomatic knee or hip osteoarthritis patients: a pilot randomized controlled trial. BMC Rheumatol 6, 46 (2022). https://doi.org/10.1186/s41927-022-00277-9 | Low prevalence of generalized anxiety |
| Mariño-Narvaez, C., Romero-Gonzalez, B., Puertas-Gonzalez, J. A., Peralta-Ramírez, M. I., & Castellote-Caballero, Y. (2025). Mindfulness-Based Stress Reduction Program for reducing anxiety and depression in hospital staff during a pandemic: A | Low prevalence of generalized anxiety |

| Reference | Reason for exclusion |
|---|---|
| randomized controlled trial. *Journal of Psychiatric Research*, *181*, 320-329. https://doi.org/https://doi.org/10.1016/j.jpsychires.2024.11.073 | |
| Marqués, M.P. (2024). Effectiveness of Mindfulness in Reducing Stress and Anxiety during Technological Change: A Randomized Controlled Trial. Romanian Journal of Applied Psychology, 26(1), https://doi.org/10.2478/rjap-2024-0005 | Modifications to MBSR exceeded those specified in the preregistered protocol. |
| McCloy, K., Hughes, C., Dunwoody, L., Marley, J., Cleland, I., Cruciani, F., & Gracey, J. (2025). Management of fatigue in gynaecological cancer: A feasibility study of an app-based exercise and mindfulness intervention. *Gynecologic Oncology Reports*, *60*, 101807. https://doi.org/https://doi.org/10.1016/j.gore.2025.101807 | Modifications to MBSR exceeded those specified in the preregistered protocol. |
| McIntyre, T.-L., Elkonin, D., de Kooker, M., & Magidson, J. F. (2018). The application of mindfulness for individuals living with HIV in South Africa: A hybrid effectiveness-implementation pilot study. *Mindfulness, 9*(3), 871–883. | Insufficient data to estimate GAD prevalence |
| McKim, R. D. (2007). *Rumination as a mediator of the effects of mindfulness: Mindfulness-based stress reduction (MBSR) with a heterogeneous community sample experiencing anxiety, depression, and/or chronic pain* (Publication No. 3290022) [Doctoral dissertation, California Institute of Integral Studies]. ProQuest Dissertations and Theses Global. | Insufficient data to estimate GAD prevalence |
| Mehta, A., Wohlgemuth, W. K., Malphurs, J. E., Claude, L. B., Gonzalez, C., & David, D. (2012). Mindfulness-based stress reduction improves total sleep time in veterans with PTSD. *Sleep, 35*, A338. | Insufficient data to estimate GAD prevalence |
| Miller, J. J., Fletcher, K., & Kabat-Zinn, J. (1995). Three-year follow-up and clinical implications of a mindfulness meditation-based stress reduction intervention in the treatment of anxiety disorders. *General Hospital Psychiatry*, *17*(3), 192-200. https://doi.org/https://doi.org/10.1016/0163-8343(95)00025-M | Low prevalence of generalized anxiety |
| Mirmahmoodi M, Mangalian P, Ahmadi A, Dehghan M. The Effect of Mindfulness-Based Stress Reduction Group Counseling on Psychological and Inflammatory Responses of the Women With Breast Cancer. Integrative Cancer Therapies. 2020;19. doi:10.1177/1534735420946819 | Modifications to MBSR exceeded those specified in the preregistered protocol. |
| Moore, R. C., Depp, C. A., Wetherell, J. L., & Lenze, E. J. (2016). Ecological momentary assessment versus standard assessment instruments for measuring mindfulness, depressed mood, and anxiety among older adults. *Journal of Psychiatric Research*, *75*, 116-123. https://doi.org/https://doi.org/10.1016/j.jpsychires.2016.01.011 | Modifications to MBSR exceeded those specified in the preregistered protocol. |

| Reference | Reason for exclusion |
|---|---|
| Morais, P., Quaresma, C., Vigário, R. et al. Electrophysiological effects of mindfulness meditation in a concentration test. Med Biol Eng Comput 59, 759–773 (2021). https://doi.org/10.1007/s11517-021-02332-y | Insufficient data to estimate GAD prevalence |
| Mousavi E, Hosseini S, Bakhtiyari M, Abasi I, Mohammadi A, Masjedi Arani A. The Effects of Mindfulness-based Stress Reduction Group Therapy on Anxiety, Depression, Stress, and the Intolerance of Uncertainty in Infertile Women. Iranian Rehabilitation Journal 2020; 18 (2) :137-144 http://irj.uswr.ac.ir/article-1-988-en.html | Insufficient data to estimate GAD prevalence |
| Mousavi F S, Marashian F S, Bakhtiarpour S. Effectiveness of Mindfulness-Based Stress Reduction and Dialectical Behavior Therapy on Pathological Worry and Difficulty in Emotion Regulation in Students With Depression Symptoms. J Research Health 2022; 12 (3) :193-202 http://jrh.gmu.ac.ir/article-1-2036-en.html | Pediatric population |
| Murano, L., Damico, V., Cossalter, L., Riggio, M., Calabresi, F., Zappia, L., & Margosio, V. (2025). The impact of Mindfulness-based stress reduction on Covid-19 survivors. A randomized controlled trial. Annali di igiene : medicina preventiva e di comunita, 37(3), 350–364. | Insufficient data to estimate GAD prevalence |
| Nasrollahi, M., Ghazanfar Pour, M., Ahmadi, A. et al. Effectiveness of mindfulness-based stress reduction on depression, anxiety, and stress of women with the early loss of pregnancy in southeast Iran: a randomized control trial. Reprod Health 19, 233 (2022). | Modifications to MBSR exceeded those specified in the preregistered protocol. |
| Nejad, F. K., Shahraki, K. A., Nejad, P. S., Moghaddam, N. K., Jahani, Y., & Divsalar, P. (2021). The influence of mindfulness-based stress reduction (MBSR) on stress, anxiety and depression due to unwanted pregnancy: A randomized clinical trial. *Journal of Preventive Medicine and Hygiene, 62*(1), E82–E88. https://doi.org/10.15167/2421-4248/jpmh2021.62.1.1691 | Insufficient data to estimate GAD prevalence |
| Nesset, M. B., Bjørngaard, J. H., Whittington, R., & Palmstierna, T. (2021). Does cognitive behavioural therapy or mindfulness-based therapy improve mental health and emotion regulation among men who perpetrate intimate partner violence? A randomised controlled trial. *International Journal of Nursing Studies*, *113*, 103795. https://doi.org/https://doi.org/10.1016/j.ijnurstu.2020.103795 | Low prevalence of generalized anxiety |
| Nice, P. R. (2008). *Mindfulness interventions in the treatment of substance and mood disorders* (Publication No. 3308482) [Doctoral dissertation, Northcentral University]. ProQuest Dissertations and Theses Global. | Modifications to MBSR exceeded those specified in the preregistered protocol. |
| Nijjar, P.S., Connett, J.E., Lindquist, R. et al. Randomized Trial of Mindfulness-Based Stress Reduction in Cardiac Patients | Low prevalence of generalized anxiety |

| Reference | Reason for exclusion |
|---|---|
| Eligible for Cardiac Rehabilitation. Sci Rep 9, 18415 (2019). https://doi.org/10.1038/s41598-019-54932-2 | |
| Norouzi, E., Naseri, A., Rezaie, L., Bender, A. M., Salari, N., & Khazaie, H. (2024). Combined mindfulness-based stress reduction and physical activity improved psychological factors and sleep quality in patients with MDD: A randomized controlled trial study. *Archives of Psychiatric Nursing*, *53*, 215-223. https://doi.org/10.1016/j.apnu.2024.10.020 | Modifications to MBSR exceeded those specified in the preregistered protocol. |
| O'Donnell, R. M. R. (2017). *Pilot RCT of mindfulness-based stress reduction (MBSR) versus progressive muscle relaxation (PMR) to reduce symptoms of distress among elderly dementia caregivers: Results at one year post-intervention* (Publication No. 10277682) [Doctoral dissertation, University of Arizona]. ProQuest Dissertations and Theses Global. | Anxiety not separately reported |
| Pagni BA, Walsh MJM, Foldes E, et al. The neural correlates of mindfulness-induced depression reduction in adults with autism spectrum disorder: A pilot study. J Neuro Res. 2020; 98: 1150–1161. https://doi.org/10.1002/jnr.24600 | Insufficient data to estimate GAD prevalence |
| Pagni, B. (2022). *Therapeutic neural mechanisms of mindfulness-based therapy in adults with autism: A functional MRI and EEG characterization* [Doctoral dissertation, Arizona State University]. | Insufficient data to estimate GAD prevalence |
| Pagni, B.A., Williams, C., Abrams, G. et al. Neurophysiological Signatures of Mindfulness-Based Stress Reduction in Adults with Autism: Putative Mechanism of Anxiety Alleviation. Mindfulness 14, 2124–2136 (2023). https://doi.org/10.1007/s12671-023-02195-4 | Insufficient data to estimate GAD prevalence |
| PagniBroc A., HillEthan, WalshMelissa J.M., DelaneyShanna, OgbeamaDestiny, MonahanLeanna, CookJames R., GuerithaultNicolas, DixonMaria V., BallardLisa, and BradenB. Blair. 2023. Distinct and shared therapeutic neural mechanisms of mindfulness-based and social support stress reduction groups in adults with autism spectrum disorder. Journal of Psychiatry and Neuroscience. 48(2): E102-E114. | Insufficient data to estimate GAD prevalence |
| Pagnini, F., Marconi, A., Tagliaferri, A., Manzoni, G.M., Gatto, R., Fabiani, V., Gragnano, G., Rossi, G., Volpato, E., Banfi, P., Palmieri, A., Graziano, F., Castelnuovo, G., Corbo, M., Molinari, E., Riva, N., Sansone, V. and Lunetta, C. (2017), Meditation training for people with amyotrophic lateral sclerosis: a randomized clinical trial. Eur J Neurol, 24: 578-586. https://doi.org/10.1111/ene.13246 | Low prevalence of generalized anxiety |
| Pal A, Mukhopadhyay P, Pal ND. An Online Mindfulness-based Intervention Decreases Stress, Anxiety, and Depression in Indian Healthcare Providers during the COVID-19 | Modifications to MBSR exceeded those specified in the preregistered protocol. |

| Reference | Reason for exclusion |
| --- | --- |
| Pandemic: A Prospective Randomized Controlled Study. J Assoc Physicians India 2024;72(12):e6-e11 | |
| Pettitt, A. K. (2024). *Digital mental health: Moderators and mechanisms of an online mental health intervention* [Doctoral dissertation, University of Oregon]. | Intervention is not MBSR |
| Poos JM, van den Berg E, Papma JM, van der Tholen FC, Seelaar H, Donker Kaat L, Kievit JA, Tibben A, van Swieten JC and Jiskoot LC (2022) Mindfulness-Based Stress Reduction in Pre-symptomatic Genetic Frontotemporal Dementia: A Pilot Study. Front. Psychiatry 13:864391. doi: 10.3389/fpsyt.2022.864391 | Low prevalence of generalized anxiety |
| Proeschold-Bell, R.J., Eagle, D.E., Tice, L.C. et al. The Selah Pilot Study of Spiritual, Mindfulness, and Stress Inoculation Practices on Stress-Related Outcomes Among United Methodist Clergy in the United States. J Relig Health 62, 2686–2710 (2023). https://doi.org/10.1007/s10943-023-01848-x | Insufficient data to estimate GAD prevalence |
| Raes, F., Griffith, J.W., Van der Gucht, K. et al. School-Based Prevention and Reduction of Depression in Adolescents: a Cluster-Randomized Controlled Trial of a Mindfulness Group Program. Mindfulness 5, 477–486 (2014). https://doi.org/10.1007/s12671-013-0202-1 | Pediatric population |
| Randolph, P., Caldera, Y., McComb, J., & Tacón, A. (2003). Mindfulness Meditation, Anxiety Reduction, and Heart Disease: A Pilot Study. Family & Community Health, 26(1), 25–33. | Insufficient data to estimate GAD prevalence |
| Reich, R. R., Lengacher, C. A., Alinat, C. B., Kip, K. E., Paterson, C., Ramesar, S.,…Park, J. (2017). Mindfulness-Based Stress Reduction in Post-treatment Breast Cancer Patients: Immediate and Sustained Effects Across Multiple Symptom Clusters. *Journal of Pain and Symptom Management*, *53*(1), 85-95. https://doi.org/10.1016/j.jpainsymman.2016.08.005 | Modifications to MBSR exceeded those specified in the preregistered protocol. |
| Reynolds, J. W. (2006). *Mechanisms of action in mindfulness-based stress reduction (MBSR), and specific factors mediating the reduction of anxiety* (Publication No. 3437809) [Doctoral dissertation, George Mason University]. ProQuest Dissertations and Theses Global. | Insufficient data to estimate GAD prevalence |
| Riley TD, Roy S, Parascando JA, et al. Mindfulness-Based Stress Reduction Live Online During the COVID-19 Pandemic: A Mixed Methods Feasibility Study. Journal of Integrative and Complementary Medicine. 2022;28(6):497-506. doi:10.1089/jicm.2021.0415 | Low prevalence of generalized anxiety |
| Robles, E., Blanco, I., Díez, G., & Vázquez, C. (2025). Mindfulness-based stress reduction for chronic pain: Enhancing psychological well-being without altering attentional biases | Modifications to MBSR exceeded those specified in the preregistered protocol. |

| Reference | Reason for exclusion |
| --- | --- |
| towards pain faces. European Journal of Pain, 29, e4714. https://doi.org/10.1002/ejp.4714 | |
| Sakki, S.E., Penttinen, H.M., Hilgert, O.M. et al. Mindfulness is associated with improved psychological well-being but no change in stress biomarkers in breast cancer survivors with depression: a single group clinical pilot study. BMC Women's Health 22, 518 (2022). | Insufficient data to estimate GAD prevalence |
| Sanilevici, M. (2021). *Mindfulness-based stress reduction (MBSR) increases mental wellbeing and emotion regulation during the first wave of the COVID-19 pandemic: A synchronous online intervention study* [Master's thesis, Interdisciplinary Center Herzliya]. | Insufficient data to estimate GAD prevalence |
| Santamaría-Peláez, M., González-Bernal, J. J., Verdes-Montenegro-Atalaya, J. C., Pérula-de Torres, L. Á., Roldán-Villalobos, A., Romero-Rodríguez, E., Hachem Salas, N., Magallón Botaya, R., González-Navarro, T. d. J., Arias-Vega, R., Valverde, F. J., Jiménez-Barrios, M., Mínguez, L. A., León-del-Barco, B., Soto-Cámara, R., & González-Santos, J. (2021). Mindfulness-Based Program for Anxiety and Depression Treatment in Healthcare Professionals: A Pilot Randomized Controlled Trial. Journal of Clinical Medicine, 10(24), 5941. https://doi.org/10.3390/jcm10245941 | Modifications to MBSR exceeded those specified in the preregistered protocol. |
| Sarebannezhad, F., Omidi, A., Zanjani, Z., Ghasemzadeh Hoseini, E., & Gholizadeh, N. (2024). The effectiveness of mindfulness-based stress reduction therapy on disease signs and symptoms and psychological distress in women with oral lichen planus. *Journal of Mazandaran University of Medical Sciences, 34*(240), 56–67. | Modifications to MBSR exceeded those specified in the preregistered protocol. |
| Sayadi, A. R., Seyed Bagheri, S. H., Khodadadi, A., & Jafari Torababadi, R. (2022). The effect of mindfulness-based stress reduction (MBSR) training on serum cortisol levels, depression, stress, and anxiety in type 2 diabetic older adults during the COVID-19 outbreak. *Journal of Medicine and Life, 15*(12), 1493–1501. https://doi.org/10.25122/jml-2021-0437 | Insufficient data to estimate GAD prevalence |
| Sen, A., Bandyopadhyay, T., Bhattacharyya, R., Saha, P. K., & Das, D. (2024). A comparative study of effect of mindfulness-based stress reduction on psychological stress & quality of life in patients of rheumatoid arthritis with waitlist control group. *Journal of the Indian Medical Association, 122*(5), 28–32. | Insufficient data to estimate GAD prevalence |
| Shapiro, S.L., Schwartz, G.E. & Bonner, G. Effects of Mindfulness-Based Stress Reduction on Medical and Premedical Students. J Behav Med 21, 581–599 (1998). https://doi.org/10.1023/A:1018700829825 | Insufficient data to estimate GAD prevalence |

| Reference | Reason for exclusion |
| --- | --- |
| Sharpe, L., Bisby, M. A., Menzies, R. E., Boyse, J. B., Richmond, B., Todd, J., Sesel, A. L., & Dear, B. F. (2025). A Tale of Two Treatments: A Randomised Controlled Trial of Mindfulness or Cognitive Behaviour Therapy Delivered Online for People with Rheumatoid Arthritis. Psychotherapy and psychosomatics, 94(2), 89–100. | Modifications to MBSR exceeded those specified in the preregistered protocol. |
| Sherr, L. J. (2010). *Moderators of the effectiveness of a mindfulness-based stress reduction intervention compared to an active control for solid organ transplant patients* (Publication No. 3422615) [Doctoral dissertation, University of Minnesota]. ProQuest Dissertations and Theses Global. | Insufficient data to estimate GAD prevalence |
| Sheybani F, Dabaghi P, Najafi S, Rajaeinejad M. Effectiveness of Mindfulness-based Stress Reduction (MBSR) on Patients With Chronic Pain: A Randomized Clinical Trial. IJPCP 2022; 28 (2) :182-195 http://ijpcp.iums.ac.ir/article-1-3307-en.html | Modifications to MBSR exceeded those specified in the preregistered protocol. |
| Sibinga, E. M. S., Perry-Parrish, C., Chung, S.-e., Johnson, S. B., Smith, M., & Ellen, J. M. (2013). School-based mindfulness instruction for urban male youth: A small randomized controlled trial. *Preventive Medicine*, *57*(6), 799-801. https://doi.org/https://doi.org/10.1016/j.ypmed.2013.08.027 | Pediatric population |
| Simpson, R., Mair, F.S. & Mercer, S.W. Mindfulness-based stress reduction for people with multiple sclerosis – a feasibility randomised controlled trial. BMC Neurol 17, 94 (2017). https://doi.org/10.1186/s12883-017-0880-8 | Low prevalence of generalized anxiety |
| Sizoo, B. B., & Kuiper, E. (2017). Cognitive behavioural therapy and mindfulness based stress reduction may be equally effective in reducing anxiety and depression in adults with autism spectrum disorders. *Research in Developmental Disabilities*, *64*, 47-55. https://doi.org/https://doi.org/10.1016/j.ridd.2017.03.004 | Modifications to MBSR exceeded those specified in the preregistered protocol. |
| Skovbjerg, S., Kolls, M., Kjærbye-Thygesen, A. et al. Mindfulness-Based Stress Reduction adapted to pregnant women with psychosocial vulnerabilities - a randomized feasibility trial in a Danish hospital-based outpatient setting. Pilot Feasibility Stud 11, 86 (2025). https://doi.org/10.1186/s40814-025-01670-0 | Insufficient data to estimate GAD prevalence |
| Smith, J. D. (2009). *Mindfulness-based stress reduction (MBSR) for women with PTSD surviving domestic violence* (Publication No. 3370129) [Doctoral dissertation, Fielding Graduate University]. ProQuest Dissertations and Theses Global. | Insufficient data to estimate GAD prevalence |
| Sobhani E, Babakhani N, Alebouyeh M R. The Effectiveness of Mindfulness-Based Stress Reduction on the Depression, Anxiety, Stress, and Pain Perception in Females with Obstructed Labour-Induced Chronic Low Back Pain. IJPCP | Insufficient data to estimate GAD prevalence |

| Reference | Reason for exclusion |
| --- | --- |
| 2019; 25 (3) :266-277 http://ijpcp.iums.ac.ir/article-1-2905-en.html | |
| Sumińska, S., & Rynkiewicz, A. (2025). Mindfulness-based stress reduction training supplemented with physiological signals from smartwatch improves mindfulness and reduces stress, but not anxiety and depression. *PLOS ONE*, *20*(4), e0322413. https://doi.org/10.1371/journal.pone.0322413 | Low prevalence of generalized anxiety |
| Tabernero, C., Gutiérrez-Domingo, T., Steca, P., Castillo-Mayén, R., Cuadrado, E., Rubio, S. J., Farhane-Medina, N. Z., & Luque, B. (2022). Effectiveness of Mindfulness and Positive Strengthening mHealth Interventions for the Promotion of Subjective Emotional Wellbeing and Management of Self-Efficacy for Chronic Cardiac Diseases. Journal of Personalized Medicine, 12(12), 1953. https://doi.org/10.3390/jpm12121953 | Modifications to MBSR exceeded those specified in the preregistered protocol. |
| Taghvaeinia, A., Karami, M. & Azizi, A. Comparison of the Effect of Dialectical Behavior Therapy, Acceptance and Commitment Therapy mindfulness-based Stress Reduction on Irritable Bowel Syndrome Symptoms, Quality of Life, Anxiety and Depression: A Pilot Randomized Controlled Trial. Psychiatr Q 95, 53–68 (2024) | Insufficient data to estimate GAD prevalence |
| Tarrasch R, Berman Z and Friedmann N (2016) Mindful Reading: Mindfulness Meditation Helps Keep Readers with Dyslexia and ADHD on the Lexical Track. Front. Psychol. 7:578. doi: 10.3389/fpsyg.2016.00578 | Insufficient data to estimate GAD prevalence |
| Tory, P. B. (2003). *A mindfulness-based stress reduction program for the treatment of anxiety* (Publication No. 3111023) [Doctoral dissertation, Hofstra University]. ProQuest Dissertations and Theses Global. | Modifications to MBSR exceeded those specified in the preregistered protocol. |
| Tran T, Finlayson M, Nalder E, Trothen T, Donnelly C. Occupational Therapist-Led Mindfulness Training Program for Older Adults Living with Early Cognitive Decline in Primary Care: A Pilot Randomized Controlled Trial. Journal of Alzheimer's Disease Reports. 2023;7(1):775-790. doi:10.3233/ADR-230006 | Insufficient data to estimate GAD prevalence |
| Trotter, M. J. (2009). *Effects of participation in a mindfulness-based stress reduction program on college students' psychological well-being* [Doctoral dissertation]. ProQuest Dissertations and Theses Global. | Anxiety not separately reported |
| Vala, M., Razmandeh, R., Rambod, K., Nasli Esfahani, E., & Ghodsi Ghasemabadi, R. (2016). Mindfulness-based stress reduction group training on depression, anxiety, stress, self-confidence and hemoglobin A1C in young women with type 2 diabetes. *Iranian Journal of Endocrinology and Metabolism, 17*(5), 382–390. | Insufficient data to estimate GAD prevalence |

| Reference | Reason for exclusion |
|---|---|
| Varur, S., & Baligar, V. P. (2024). *Evaluating the effectiveness of MBSR on engineering students using PSS & STAI* (pp. 189–194). Research in Engineering Education Network. https://doi.org/10.52202/073963-0024 | Insufficient data to estimate GAD prevalence |
| Vinesett AL, Whaley RR, Woods-Giscombe C, et al. Modified African Ngoma Healing Ceremony for Stress Reduction: A Pilot Study. The Journal of Alternative and Complementary Medicine: Paradigm, Practice, and Policy Advancing Integrative Health. 2017;23(10):800-804. doi:10.1089/acm.2016.0410 | Anxiety not separately reported |
| Vøllestad, J., Sivertsen, B., & Nielsen, G. H. (2011). Mindfulness-based stress reduction for patients with anxiety disorders: Evaluation in a randomized controlled trial. *Behaviour Research and Therapy*, *49*(4), 281-288. https://doi.org/https://doi.org/10.1016/j.brat.2011.01.007 | Low prevalence of generalized anxiety |
| Vyas, A. P. (2022). *Effects of mindfulness and stress management on neurocardiovascular and psychological outcomes* [Doctoral dissertation, Michigan Technological University]. | Low prevalence of generalized anxiety |
| Watson-Singleton, N. N., & Pennefather, J. (2024). Using a Randomized Clinical Trial to Test the Efficacy of a Culturally Responsive Mobile Health Application in African Americans. *Behavior Therapy*, *55*(4), 813-824. https://doi.org/https://doi.org/10.1016/j.beth.2023.12.002 | Modifications to MBSR exceeded those specified in the preregistered protocol. |
| Wells, R.E., Burch, R., Paulsen, R.H., Wayne, P.M., Houle, T.T. and Loder, E. (2014), Meditation for Migraines: A Pilot Randomized Controlled Trial. Headache: The Journal of Head and Face Pain, 54: 1484-1495. https://doi.org/10.1111/head.12420 | Insufficient data to estimate GAD prevalence |
| Wetherell, J. L., Hershey, T., Hickman, S., Tate, S. R., Dixon, D., Bower, E. S., & Lenze, E. J. (2017). Mindfulness-based stress reduction for older adults with stress disorders and neurocognitive difficulties: A randomized controlled trial. *The Journal of Clinical Psychiatry, 78*(7), e734–e743. https://doi.org/10.4088/JCP.16m10947 | Insufficient data to estimate GAD prevalence |
| Whitebird, R. R., Kreitzer, M. J., Vazquez-Benitez, G., & Enstad, C. J. (2018). Reducing diabetes distress and improving self-management with mindfulness. Social Work in Health Care, 57(1), 48–65. https://doi.org/10.1080/00981389.2017.1388898 | Insufficient data to estimate GAD prevalence |
| Wu Xuemei, Hu Jiannv, Xie Lihua, et al. Observation on the effect of mindfulness-based stress reduction intervention on improving negative emotions and sleep quality in HIV-infected | Insufficient data to estimate GAD prevalence |

| Reference | Reason for exclusion |
|---|---|
| pregnant women [J]. Chinese Journal of Clinical Infectious Diseases, 2023, 16(03): 225-229. DOI: 10.3760/cma.j.issn.1674-2397.2023.03.011 | |
| Wu, M., Fan, C., Zhao, X., Zhou, J., Liu, H., Li, H., Zhan, X., & Jiang, Z. (2024). The effects of mindfulness-based stress reduction in mental and physical health: Is mindful breathing key? *Mindfulness, 15*(10), 2582–2594. https://doi.org/10.1007/s12671-024-02456-w | Insufficient data to estimate GAD prevalence |
| Wu, Y., Ban, Y., Pan, G. et al. Effects of online mindfulness-based stress reduction training on depression and anxiety symptoms among psychiatric healthcare workers in a randomized controlled trial: the mediating role of emotional suppression. BMC Psychiatry 25, 577 (2025). | Modifications to MBSR exceeded those specified in the preregistered protocol. |
| Würtzen, H., Dalton, S. O., Elsass, P., Sumbundu, A. D., Steding-Jensen, M., Karlsen, R. V.,…Johansen, C. (2013). Mindfulness significantly reduces self-reported levels of anxiety and depression: Results of a randomised controlled trial among 336 Danish women treated for stage I–III breast cancer. *European Journal of Cancer*, *49*(6), 1365-1373. https://doi.org/10.1016/j.ejca.2012.10.030 | Insufficient data to estimate GAD prevalence |
| Yay-Pence, A., Aslan, S., Coldur, M., Atalay, Z., Beba, B., Coskun-Sayin, C.,…Ekmekci-Ertek, I. (2025). Online mindfulness-based vs. cognitive-behavioural stress reduction for medical students: A randomized controlled trial. *Psychiatry Research*, *351*, 116632. https://doi.org/https://doi.org/10.1016/j.psychres.2025.116632 | Low prevalence of generalized anxiety |
| Zhang, J. Y., Zhou, Y. Q., Feng, Z. W., Fan, Y. N., Zeng, G. C., & Wei, L. (2017). Randomized controlled trial of mindfulness-based stress reduction (MBSR) on posttraumatic growth of Chinese breast cancer survivors. Psychology, Health & Medicine, 22(1), 94–109. https://doi.org/10.1080/13548506.2016.1146405 | Insufficient data to estimate GAD prevalence |
| Zhang, L. (2024). Intervention Treatment for Anxiety and Depression-Induced Emotional Disorders in College Students: Mindfulness-Based Stress Reduction. *Journal of Engineering and Science in Medical Diagnostics and Therapy*, *7*(3). https://doi.org/10.1115/1.4064345 | Low prevalence of generalized anxiety |
| Zhou Y, Tang Z-M, Mei L, Yang X-C, Zheng H-L, Liao B-Z and Yu X-Y (2025) Effect of mindfulness group therapy on maternal psychological distress and perinatal outcomes in twin pregnancy: a randomized controlled trial. Front. Psychiatry 16:1572633. doi: 10.3389/fpsyt.2025.1572633 | Modifications to MBSR exceeded those specified in the preregistered protocol. |

#### Appendix 4. Exclusion of Study: Data Integrity Assessment

One study was excluded from the meta-analysis after routine pre-synthesis checks identified features that, taken together, prevented reliable inclusion in our pooled estimates (8). When effect size was calculated from the reported raw means and baseline SDs, the resulting estimate differed from the estimate derived from the reported ANOVA MS error; the control group's pre- and post-treatment means on one measure were also reported as identical. Preliminary influence diagnostics further identified the study as a statistical outlier whose inclusion substantially increased heterogeneity and shifted the pooled effect, potentially biasing towards a larger estimated effect of MBSR. Given the difficulty of resolving these features without access to the original data, and the outsized influence a single such study would exert on our synthesis (biasing towards an effect of MBSR), we opted for the more conservative choice of exclusion, which biases findings toward the null rather than away from it.

#### Appendix 5: Supplemental Figures and Tables

**Figure S1:** PRISMA Flowchart of Study Selection

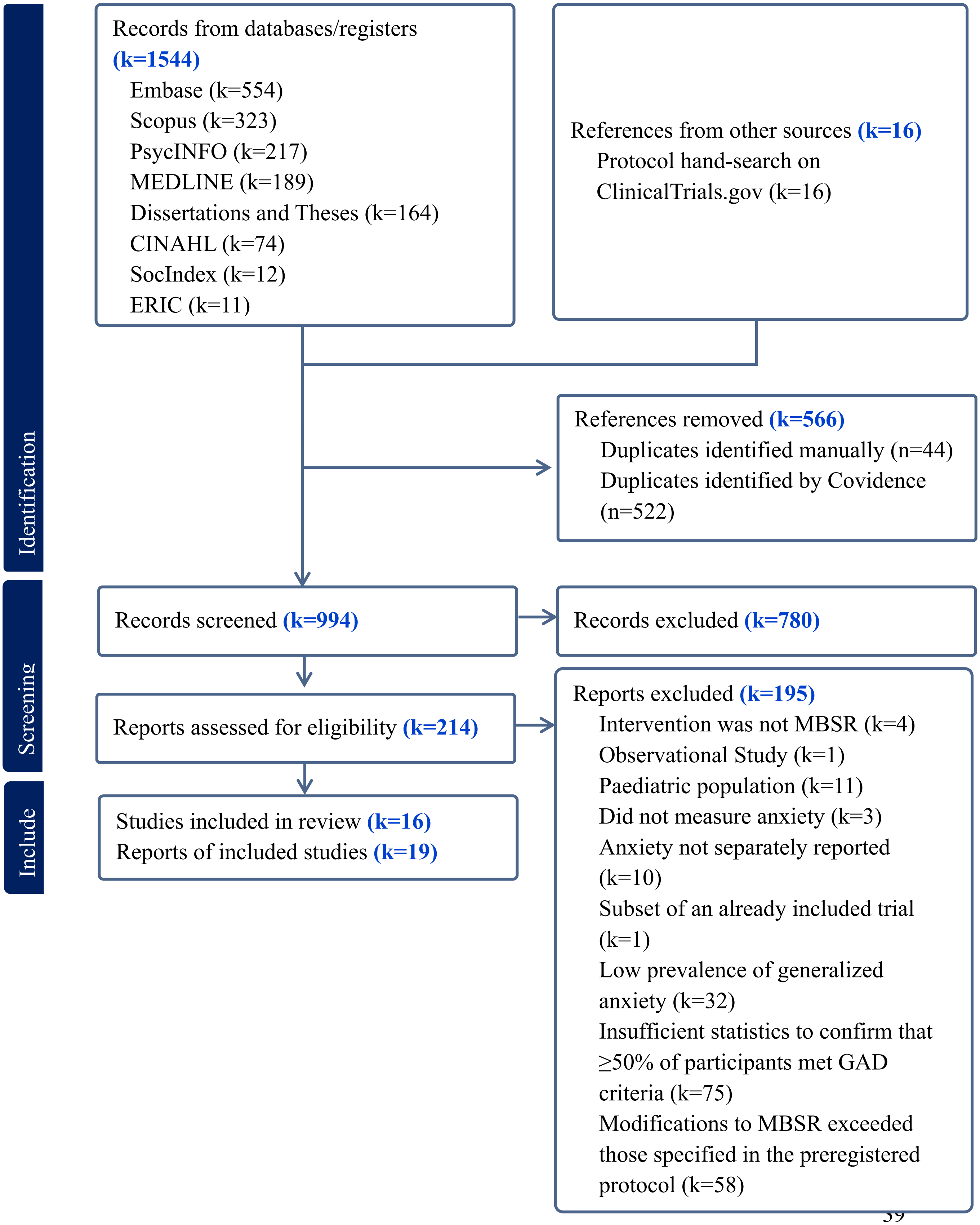

**Figure S2.** Forest plot of longer-term between-group differences in anxiety symptoms (Hedges' g), stratified by comparator type. For each study, change scores (long-term follow-up minus baseline) were computed for the MBSR and control arms, standardized by the pooled SD, and corrected for small-sample bias. Effects were pooled within each comparator stratum using a random-effects model. Blue diamonds represent summary effects for General Active Comparators and First-Line Treatment for Anxiety.

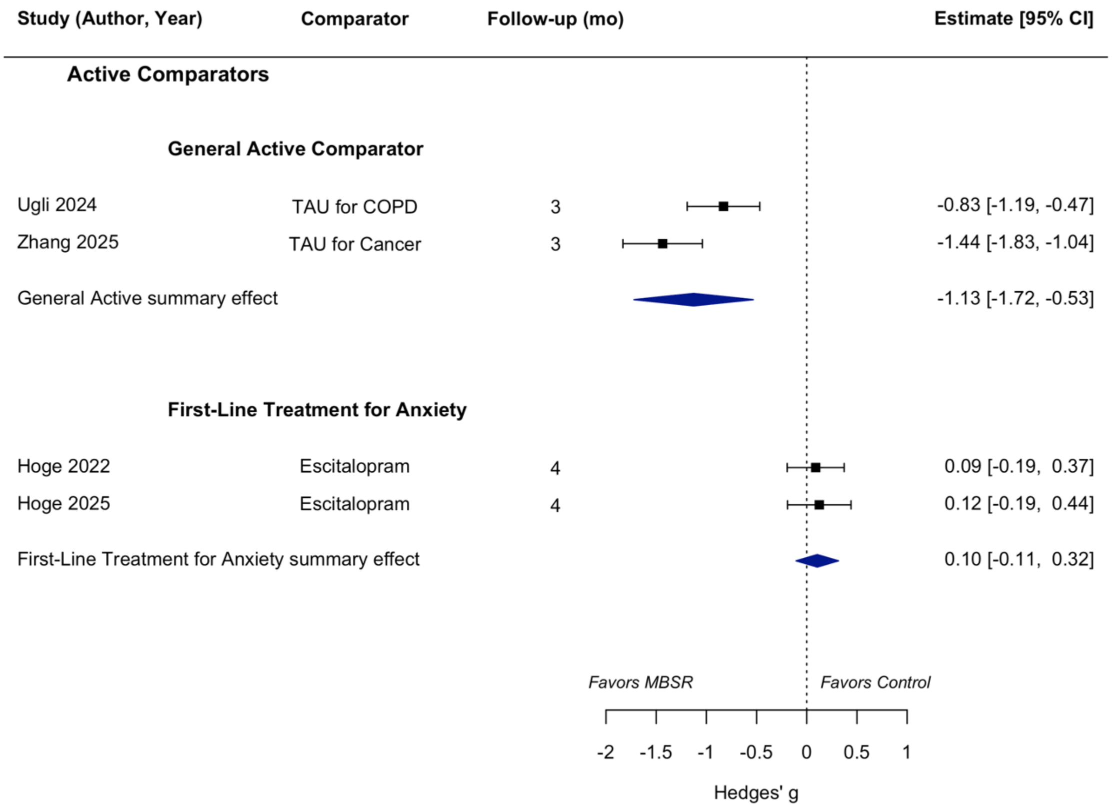

**Figure S3.** Random-effects meta-analysis of within-group change in anxiety symptoms from baseline to immediate post-treatment in the MBSR arms only. For each trial, we computed the standardized mean change as the difference between pre- and post-treatment scores divided by the baseline standard deviation (Becker's method). A pre–post correlation of 0.7 was assumed to derive the variance for all studies, with Hedges' small-sample correction applied. Negative values indicate greater improvement in anxiety symptoms (reductions from baseline), while values closer to zero indicate less improvement. Results are subgrouped by study design (RCT, Quasi-Experimental Study, and SAT).

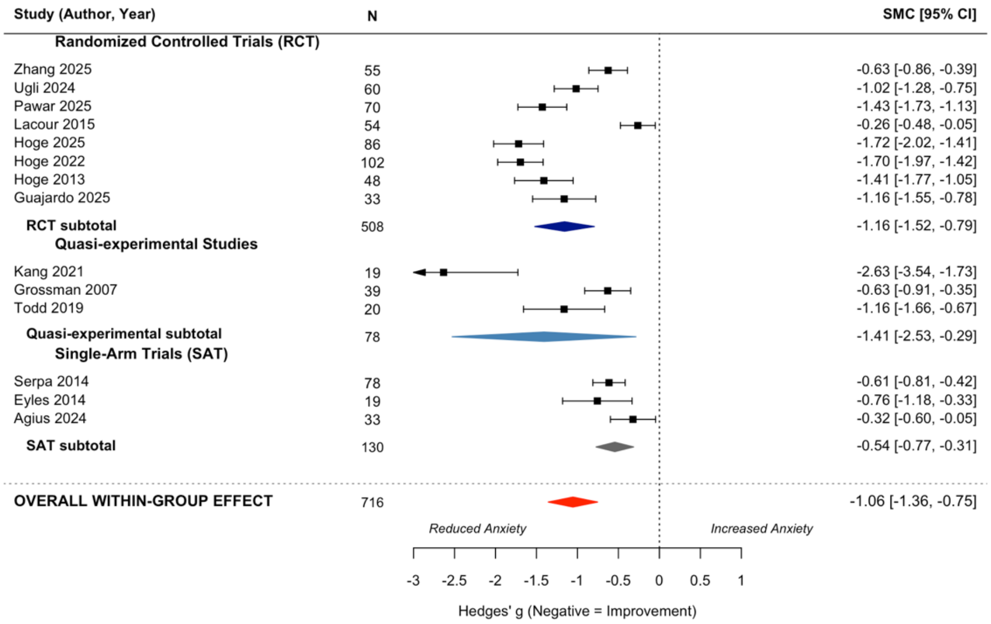

**Figure S4.** Random-effects meta-analysis of within-group change in anxiety symptoms from baseline to long-term follow-up (range: 3–36 months) in the MBSR arms only. For each trial, we computed the standardized mean change as the difference between baseline and long-term follow-up scores divided by the baseline standard deviation (Becker's method). A pre–post correlation of 0.7 was assumed to derive the variance for all studies, and Hedges' small-sample correction was applied to obtain Hedges' g. Negative values indicate sustained improvement in anxiety symptoms, whereas values closer to zero indicate less long-term benefit. The "Follow-up (mo)" column indicates the timing of the final assessment for each study (in months).

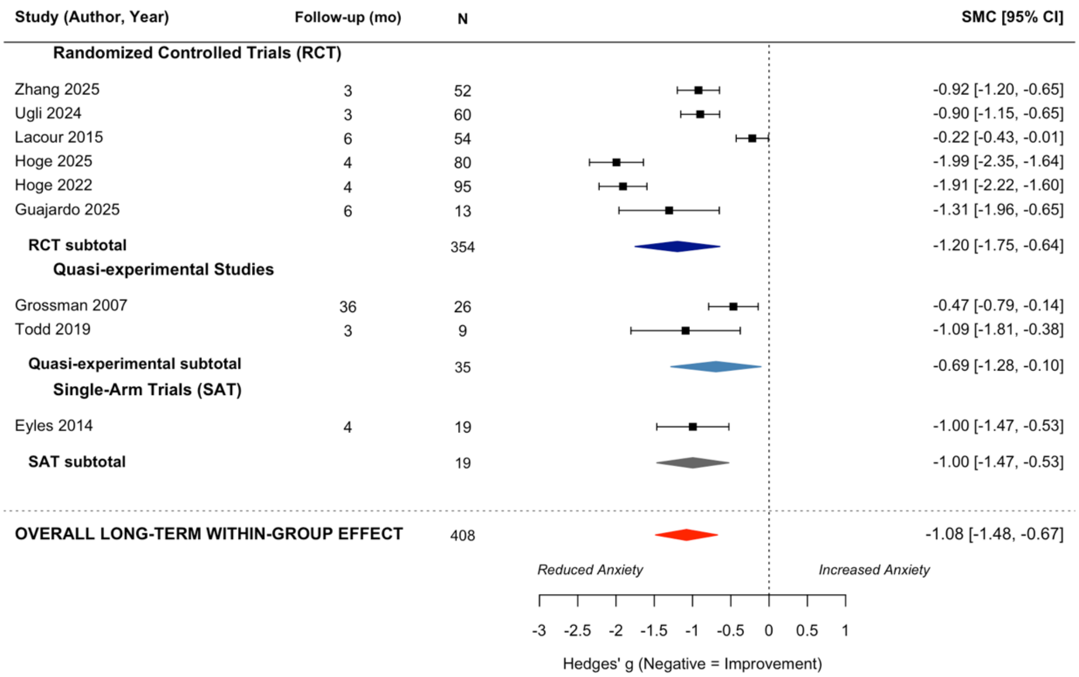

**Table S1**: Pre-registered Moderator Analysis

| Moderator Variable | | $M\ g$ (95% CI) | $k$ | Model Statistics | | | | |
|---|---|---|---|---|---|---|---|---|
| | | | | $B$ | $P$ | $Q_{model}$ | $Q_{residual}$ | $I^2$ |
| *Within-Group Comparisons* | | | | | | | | |
| Study Design | | | | +0.253 | 0.4180 | 0.656 | 143.194 | 92.7% |
| | Non-RCT | −0.905 (−1.376, −0.435) | 6 | | | | | |
| | RCT (Ref) | −1.158 (−1.548, −0.768) | 8 | | | | | |
| Risk of Bias (RCT Only)‡ | | | | +0.823 | 0.0520 | 5.911 | 49.309 | 88.87% |
| | High Risk of Bias | −0.884 (−1.250, −0.518) | 5 | | | | | |
| | Some Concerns | −1.410 (−2.258, −0.562) | 1 | | | | | |
| | Low Risk of Bias (Ref) | −1.707 (−2.288, −1.126) | 2 | | | | | |
| Conflict of Interest | | | | −0.373 | 0.2692 | 1.221 | 102.211 | 92.93% |
| | Yes | −1.245 (−1.711, −0.780) | 6 | | | | | |
| | No (Ref) | −0.872 (−1.343, −0.401) | 6 | | | | | |
| Study Population | | | | +0.358 | 0.3335 | 0.935 | 139.701 | 92.9% |
| | Mixed Anxiety Disorders | −0.977 (−1.315, −0.640) | 11 | | | | | |
| | GAD only (Ref) | −1.335 (−1.978, −0.693) | 3 | | | | | |
| Curriculum | | | | +0.682** | 0.0092 | 6.786 | 106.461 | 89.19% |
| | Modified MBSR Curriculum | −0.611 (−1.019, −0.203) | 5 | | | | | |
| | vs Standard (Ref) | −1.293 (−1.606, −0.981) | 9 | | | | | |
| Eligibility and outcome assessment† | | | | +0.737* | 0.0122 | 6.275 | 81.491 | 88.67% |

**Table S1**: Pre-registered Moderator Analysis

| Moderator Variable | | $M\ g$ (95% CI) | $k$ | Model Statistics | | | | |
|---|---|---|---|---|---|---|---|---|
| | | | | $B$ | $P$ | $Q_{model}$ | $Q_{residual}$ | $I^2$ |
| | Scale cutoff, self-report | −0.875 (−1.146, −0.603) | 11 | | | | | |
| | Structured interview, clinician-rated (Ref) | −1.612 (−2.120, −1.103) | 3 | | | | | |
| Format | | | | −0.821* | 0.0178 | 5.611 | 126.564 | 90.9% |
| | Synchronous Online Delivery | −1.714 (−2.327, −1.101) | 3 | | | | | |
| | In-person (Ref) | −0.893 (−1.186, −0.600) | 11 | | | | | |
| Mean Age (years) | | | 13 | +0.025 | 0.1391 | 2.188 | 130.605 | 91.85% |
| | At youngest (33 y) | −1.402 (−1.916, −0.888) | | | | | | |
| | At median (45.1 y) | −1.103 (−1.407, −0.799) | | | | | | |
| | At oldest (63 y) | −0.660 (−1.304, −0.016) | | | | | | |
| Gender (% female in sample) | | | 13 | −0.005 | 0.3673 | 0.813 | 148.514 | 93.29% |
| | At fewest female (11%) | −0.736 (−1.452, −0.021) | | | | | | |
| | At median (76%) | −1.074 (−1.416, −0.732) | | | | | | |
| | At all female (100%) | −1.199 (−1.692, −0.705) | | | | | | |
| *Between-Group Comparisons* | | | | | | | | |
| Type of Control Group | | | | +0.423 | 0.3122 | 1.021 | 62.811 | 89.86% |
| | Active Control | −0.456 (−0.953, +0.041) | 5 | | | | | |
| | Inactive Control (Ref) | −0.879 (−1.531, −0.227) | 3 | | | | | |
| Risk of Bias‡ | | | | −1.016** | 0.0051 | 10.572 | 21.279 | 77.43% |

**Table S1**: Pre-registered Moderator Analysis

| Moderator Variable | | $M$ $g$ (95% CI) | $k$ | Model Statistics | | | | |
|---|---|---|---|---|---|---|---|---|
| | | | | $B$ | $P$ | $Q_{model}$ | $Q_{residual}$ | $I^2$ |
| | High Risk of Bias | −0.929 (−1.272, −0.586) | 5 | | | | | |
| | Some Concerns | −0.521 (−1.301, +0.260) | 1 | | | | | |
| | Low Risk of Bias (Ref) | +0.087 (−0.422, +0.597) | 2 | | | | | |
| Conflict of Interest | | | | +0.530 | 0.0898 | 2.878 | 22.927 | 79.79% |
| | Yes | −0.244 (−0.644, +0.157) | 4 | | | | | |
| | No (Ref) | −0.774 (−1.238, −0.310) | 3 | | | | | |
| Study Population | | | | +0.528 | 0.1858 | 1.751 | 55.417 | 88.85% |
| | Mixed Anxiety Disorders | −0.419 (−0.890, +0.052) | 5 | | | | | |
| | GAD only (Ref) | −0.947 (−1.570, −0.323) | 3 | | | | | |
| Curriculum | | | | −0.167 | 0.7086 | 0.140 | 76.619 | 91.22% |
| | Modified MBSR Curriculum | −0.719 (−1.417, −0.020) | 3 | | | | | |
| | vs Standard (Ref) | −0.551 (−1.083, −0.019) | 5 | | | | | |
| Eligibility and outcome assessment† | | | | −0.832** | 0.0046 | 8.018 | 27.944 | 79.33% |
| | Scale cutoff, self-report | −0.928 (−1.288, −0.568) | 5 | | | | | |
| | Structured interview, clinician-rated (Ref) | −0.096 (−0.546, +0.353) | 3 | | | | | |
| Format | | | | +0.387 | 0.4204 | 0.649 | 70.498 | 90.17% |
| | Synchronous Online Delivery | −0.320 (−1.138, +0.498) | 2 | | | | | |
| | In-person (Ref) | −0.707 (−1.175, −0.240) | 6 | | | | | |

**Table S1**: Pre-registered Moderator Analysis

| Moderator Variable | $M$ $g$ (95% CI) | $k$ | Model Statistics | | | | |
|---|---|---|---|---|---|---|---|
| | | | $B$ | $P$ | $Q_{model}$ | $Q_{residual}$ | $I^2$ |
| Mean Age (years) | | 7 | −0.028 | 0.2184 | 1.515 | 49.995 | 89.44% |
| At youngest (33 y) | −0.253 (−0.881, +0.375) | | | | | | |
| At median (41 y) | −0.478 (−0.909, −0.046) | | | | | | |
| At oldest (63 y) | −1.096 (−2.063, −0.129) | | | | | | |
| Gender (% female in sample) | | 7 | +0.009 | 0.2205 | 1.501 | 29.822 | 83.8% |
| At fewest female (25%) | −0.826 (−1.480, −0.172) | | | | | | |
| At median (73%) | −0.397 (−0.753, −0.041) | | | | | | |
| At all female (100%) | −0.156 (−0.764, +0.453) | | | | | | |

*Note*. Between-group effect sizes are computed as MBSR minus control, so negative *g* favors MBSR; within-group effect sizes reflect pre-to-post change, so negative *g* indicates anxiety reduction. Each moderator is reported as a header row giving the moderator test — *B* = the random-effects meta-regression coefficient contrasting the listed category with its reference category (for continuous moderators, the change in g per one-unit increase), where a negative B indicates that the listed category is associated with greater anxiety improvement relative to its reference; *P* and $Q_{model}$= omnibus test of whether the moderator explains between-study variance; $Q_{residual}$ = residual heterogeneity remaining after accounting for the moderator; $I^2$ = percentage of residual variance attributable to true between-study heterogeneity — followed by indented rows giving the model-estimated mean effect at each level of that moderator: *M g* (95% CI) = pooled Hedges' *g* with its 95% confidence interval, and *k* = number of effects contributing to that level. Reference categories are marked "(Ref)". For continuous moderators (mean age, % female), the indented rows give the model-predicted mean *g* (and 95% CI) at the lowest, median, and highest observed values; because these studies are not divided into categories, k for these models is shown on the header row.  Category means are model-based estimates from the same meta-regression that produced *B* and are therefore conditional on that model.  *P < 0.05.  **P < 0.01.

† In the pooled studies, eligibility method and outcome source coincided: the three trials with clinician-rated outcomes were the only ones that established eligibility by structured interview.

‡ Risk of bias (RoB 2) has three levels, and the model estimates all three, so the per-level k values sum to the total number of trials. *B* reports the high-versus-low contrast only, and $Q_{model}$ is the joint 2-df omnibus test. Because a single trial was rated "some concerns" (k = 1), that level's mean reflects one study and should be interpreted with caution.

**Figure S5:** Funnel plots assessing small-studies effects for the between-group (A) and within-group (B) meta-analyses. The x-axis displays Hedges' *g* and the y-axis displays its standard error; the vertical dashed line marks the pooled random-effects estimate. The diagonal dashed lines demarcate the 95% pseudo-confidence interval region; under homogeneity, 95% of studies would be expected to fall within this boundary. **(A)**: between-group short-term effects (MBSR vs. control); symbols and colors distinguish comparator type (see inset legend); negative values favor MBSR. **(B)**: within-group pre-to-post change in MBSR arms only; negative values indicate anxiety reduction from baseline.

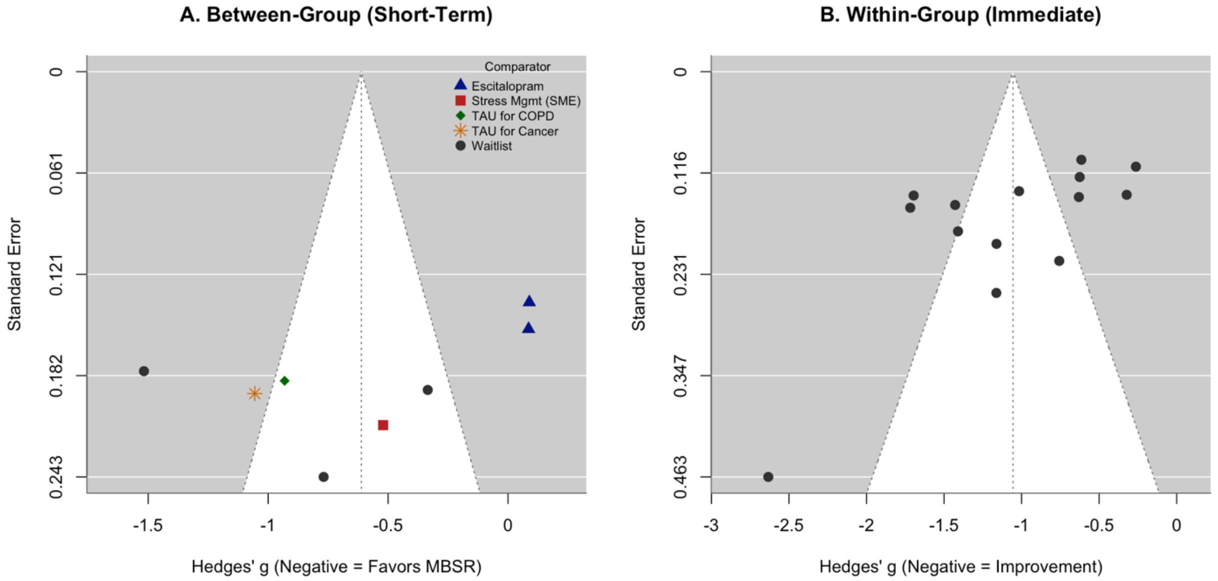

**Table S2:** Funding Sources for Included Studies

| Study | Reported Funding Source(s) | Funding Status |
|---|---|---|
| **Agius 2024** | Region Stockholm ALF agreement (FoUI-960703); Promobilia Foundation. | Reported |
| **Berghmans 2012** | Not mentioned by study authors. | Not Reported |
| **Eyles 2014** | National Institute for Health Research (NIHR), Research for Patient Benefit (Grant PB-PG-0408-16155). | Reported |
| **Grossman 2007** | HRCA Research and Training Institute; YeTaDel Foundation. | Reported |
| **Guajardo 2025** | The authors explicitly stated that "no funds, grants, or other support were received". | Reported (None) |
| **Hoge 2013** | NIH National Center on Complementary and Alternative Medicine (Grant K23AT4432); Highland Street Foundation; NIH Harvard Clinical and Translational Science Center. | Reported |
| **Hoge 2015** | NIH National Center on Complementary and Alternative Medicine (Grant K23AT4432); Highland Street Foundation. | Reported |
| **Hoge 2022** | Patient-Centered Outcomes Research Institute (PCORI; CER-2017C1–6522); NYU Innovation Fund (partial support for treatments). | Reported |
| **Hoge 2024** | Intramural Research Program of the National Institute of Mental Health (Grant ZIAMH002798); internal departmental funds. | Reported |
| **Hoge 2025** | Patient-Centered Outcomes Research Institute (PCORI; CER-2017C1–6522); private donor; NYU Innovation Fund (partial support for treatments). | Reported |
| **Hu 2024** | Patient-Centered Outcomes Research Institute (PCORI; CER-2017C1-6522); NYU Innovation Fund. | Reported |
| **Kang 2021** | National Natural Scientific Foundation of China (Grants 81660235 and 81971288); Tongji University COVID-19 special fund; NIH (K23AT011173). | Reported |
| **La Cour 2015** | TrygFonden; Axel Muusfeldts Fond; Fabrikant Mads Clausens Fond; Fonden af 1870. | Reported |
| **Majid 2012** | The authors explicitly declared that the study was "not support by any financial source". | Reported (None) |
| **Pawar 2025** | Not mentioned by study authors. | Not Reported |
| **Serpa 2014** | VA Office of Patient Centered Care & Cultural Transformation. | Reported |
| **Todd 2019** | National Union of Teachers (NUT); DECIPHer (UKCRC); British Heart Foundation; Cancer Research UK; ESRC; MRC; Welsh Government; Wellcome Trust; Austin Bailey Foundation; NCPHWR. | Reported |
| **Ugli 2024** | Not mentioned in the provided excerpts. | Not Reported |
| **Zhang 2025** | Henan Medical Science and Technology Research Project Fund (Grants LHGJ20230512 and 222102310457). | Reported |

### Appendix 6. Adverse Events: Study-Level Detail

Of the 16 included studies, 5 (31%) explicitly documented adverse events: three reported specific occurrences and two reported none.

*la Cour et al. (2015)*: Passive surveillance identified four adverse events in the MBSR group and none in the control group. Events consisted of temporary anger toward the clinical condition (n=2) and increased anxiety (n=2).

*Hoge et al. (2013)*: One event of mild muscle soreness (2%) was reported in the MBSR arm and one event of sleep disruption (2%) in the control arm.

*Hoge et al. (2023) — MBSR versus escitalopram*: Using active clinician-led safety monitoring, 21 MBSR participants (15%) and 110 escitalopram participants (79%) reported at least one study-related adverse event. Zero MBSR participants and 10 escitalopram participants (8%) discontinued treatment due to adverse events. The most common adverse events in this trial were transient increased anxiety (MBSR: 11%, n=13; escitalopram: 5%, n=7), insomnia/sleep disturbance (MBSR: <1%; escitalopram: 41%), nausea (MBSR: <1%; escitalopram: 35%), fatigue (MBSR: 2%; escitalopram: 26%), headache (MBSR: 2%; escitalopram: 18%), and somnolence (MBSR: 0%; escitalopram: 14%). Several additional events — including anorgasmia/delayed orgasm, abnormal dreaming, decreased appetite, jitteriness, decreased libido, dizziness, and increased sweating — each affected 6–11% of the escitalopram arm, while none exceeded 1% in the MBSR arm.
